# The Cultural Evolution of Vaccine Hesitancy: Modeling the Interaction between Beliefs and Behaviors

**DOI:** 10.1101/2022.05.26.22275604

**Authors:** Kerri-Ann Anderson, Nicole Creanza

## Abstract

Health perceptions and health-related behaviors can change at the population level as cultures evolve. In the last decade, despite the proven efficacy of vaccines, the developed world has seen a resurgence of vaccine-preventable diseases (VPDs) such as measles, pertussis, and polio. Vaccine hesitancy, an individual attitude influenced by historical, political, and socio-cultural forces, is believed to be a primary factor responsible for decreasing vaccine coverage, thereby increasing the risk and occurrence of VPD outbreaks. In recent years, mathematical models of disease dynamics have begun to incorporate aspects of human behavior, however they do not address how beliefs and motivations influence these health behaviors. Here, using a mathematical modeling framework, we explore the effects of cultural evolution on vaccine hesitancy and vaccination behavior. With this model, we shed light on facets of cultural evolution (vertical and oblique transmission, homophily, etc.) that promote the spread of vaccine hesitancy, ultimately affecting levels of vaccination coverage and VPD outbreak risk in a population. In addition, we present our model as a generalizable framework for exploring cultural evolution when humans’ beliefs influence, but do not strictly dictate, their behaviors. This model offers a means of exploring how parents’ potentially conflicting beliefs and cultural traits could affect their children’s health and fitness. We show that vaccine confidence and vaccine-conferred benefits can both be driving forces of vaccine coverage. We also demonstrate that an assortative preference among vaccine-hesitant individuals can lead to increased vaccine hesitancy and lower vaccine coverage.

## 1. Introduction

Niche construction is a process in which organisms modify their local environment, thus altering selection pressures on themselves and the other organisms in that environment [1,2]. In *cultural* niche construction, humans modify their cultural environments—such as their beliefs, behaviors, preferences, and social contacts—in ways that subsequently alter evolutionary pressures on themselves and/or their culture [2]. Mathematical models of niche construction have traditionally been used in a biological and ecological context. More recently, this type of model has been expanded to explain the evolution of cultural behaviors, with applications to religion, fertility, and the evolution of large-scale human conflict [2–7]. Cultural niche construction theory recognizes that human evolution is in part directed by human behaviors. As such, using a niche construction framework allows for the exploration of a broader range of complex feedback scenarios resulting from selection pressures that may be caused by—or act upon—non-genetic traits. It may also provide insight into how otherwise deleterious traits become beneficial in certain environments and thus spread in a population [1]. Here, we propose a cultural niche construction model of the interactions between beliefs and behaviors, in which an individual’s beliefs influence their behaviors, and these belief-behavior interactions can be affected by and shape the broader cultural landscape. We apply this model to the interactions between vaccine-related beliefs, such as vaccine hesitancy of individual parents, and vaccination behaviors, such as a pair of parents vaccinating their offspring. Modeling the belief-behavior interactions underlying vaccination coverage using a cultural niche construction framework allows us to better understand how vaccination “cultures” are formed and how they can be transformed to promote public health.

Understanding vaccination behaviors is a crucial aspect of preventing infectious disease outbreaks. The implementation of childhood vaccination policies has led to the eradication of smallpox and the elimination of poliomyelitis (polio) in the United States [8–10]. The high efficacy of the measles vaccine, combined with wide vaccine acceptance in developed countries, had resulted in measles previously being targeted for elimination by 2020 [11]. However, over the past decade, there has been a resurgence of vaccine-preventable diseases (VPDs) in developed countries despite the safety and efficacy of vaccines and high overall childhood vaccination rates [12–15]. Vaccine hesitancy, named one of the World Health Organization’s ten threats to global health in 2019 [16], is believed to be responsible for decreasing vaccination coverage and thus increasing the risk of vaccine-preventable disease outbreaks worldwide [17]. Vaccine hesitancy is a complex and context-specific individual attitude influenced by multiple factors, such as complacency (the belief that vaccination is unnecessary when the perceived risk of VPDs is low), convenience (the accessibility and affordability of vaccines), and confidence (the level of trust in the efficacy and safety of the vaccine, and in the healthcare system) [15,18]. Additionally, anti-vaccine sentiments are still on the rise despite well-documented vaccine efficacy and safety, including numerous studies debunking the spurious connection between vaccines and autism [19] and other anti-vaccination arguments [20]. The spread of these sentiments and disease outbreak risk are further exacerbated by homophily—the tendency of individuals to choose social contacts and mates who are similar to themselves [6,7,21,22]. Network-based simulations suggest that individuals with similar vaccine-hesitant opinions form groups that are more susceptible to vaccine-preventable diseases, impeding the attainment of herd immunity and substantially increasing the likelihood of disease outbreak in these clusters [23].

Even though some epidemiological models have begun to include aspects of human behavior (e.g [24–26]), these models do not typically incorporate the effects of population beliefs and changing cultural landscapes on disease transmission. For example, established epidemiological models such as the Susceptible-Infected-Recovered (SIR) model have been modified to include a vaccination component that can be useful in determining the intensity of intervention needed to address an epidemic [27]; however these models do not generally address fluctuations in vaccination rates or lower-than-expected rates of adoption based on cultural factors. One notable exception is a recent study in which an SIR model of a contagious disease was paired with another SIR model in which vaccine hesitancy is treated as a “cultural contagion”; this model showed that the spread of anti-vaccine sentiment could cause epidemics that would otherwise not have occurred [28]. However, it is still important that we understand how parents’ beliefs, which may differ from one another, interact with their perceptions of the relative risks of disease and vaccines to shape the decision to vaccinate their children, which in turn affects the future risk of vaccine-preventable disease outbreaks. Indeed, belief systems can act as the main barrier to vaccination, as opposed to lack of vaccine access, particularly in wealthier countries [23,29]. For example, increasing rates of non-medical exemption from vaccines (exemption on the basis of religious, philosophical, and personal beliefs), have been observed in the United States [30,31]. Without these considerations, models commonly used in public health may be misleading; thus, understanding and incorporating the underlying health cultures and their evolution, including the interplay between beliefs and behaviors, will allow us to build more comprehensive and representative models of vaccination dynamics and better support public health efforts.

In this study, we model the development and spread of vaccine hesitancy and childhood vaccination through a cultural evolution framework, incorporating the transmission of vaccine attitudes both from parents and from the community. We aim to assess the dynamic interactions between beliefs (shaped by social interactions) and behaviors (influenced by these beliefs). Using vaccine hesitancy and vaccination behaviors as a focal example of belief-behavior interactions, we explore the situations in which vaccine hesitancy is most likely to spread, potentially reducing childhood vaccination rates and leading to an increase in vaccine-preventable disease outbreaks. In addition, we consider that the perception of the relative risks of a disease and its preventive vaccine can fluctuate based on the prevalence of vaccination [26], such that the population’s vaccination coverage can influence the decision to vaccinate one’s children. Finally, we take into account that the decision to vaccinate a child is often the joint consideration of two individuals who might have different vaccine attitudes, and we further incorporate homophily (assortative mating) to understand how social subcultures might influence parental behaviors. Overall, we propose that a generalizable modeling framework for belief-behavior interactions can help inform public health strategies by improving our understanding of the cultural dynamics of vaccine hesitancy.

## 2. Methods

To model the evolution of vaccine beliefs and behaviors, we build on the cultural niche construction framework of [6] to assess the effects of vaccine attitudes on vaccination behaviors and on the resulting vaccination culture. We use this adapted model to explore how vaccination patterns evolve in a population when a cultural trait, such as vaccine hesitancy, can influence but not perfectly predict a behavior, such as vaccinating one’s children.

We consider two cultural traits: **V**, a vaccination trait, and **A**, a vaccine attitude trait. Each trait has two possible states, V^**+**^ (vaccinated) or V^**−**^ (unvaccinated) and A^+^ (vaccine confident) or A^−^ (vaccine hesitant), respectively. Thus, there are four possible phenotypes: V^+^A^+^ (type 1: vaccinated and confident), V^+^A^−^ (type 2: vaccinated and hesitant), V^−^A^+^ (type 3: unvaccinated and confident), and V^−^A^−^ (type 4: unvaccinated and hesitant), whose population frequencies are denoted by *x*_1_, *x*_2_, *x*_3_, and *x*_4_, respectively, with 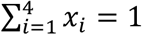.

The attitude trait (**A**) can influence the dynamics of the vaccination trait (**V**) in two ways: by affecting the likelihood that couples vaccinate their offspring, and by determining with whom each adult will preferentially pair in assortative interactions. The state of the vaccine attitude trait (**A**) informs the value of an assortative mating parameter (*α*_*k*_), which measures the departure from random mating. We define a ‘choosing parent’, arbitrarily, as the first member of each mating pair. The choosing parent’s **A** state dictates the level of assortative mating, that is, the degree to which an individual of a given **A** state will preferentially mate with another individual of the same state, expressed by parameters *α*_*k*_ where *k* = {1, 2} and 0≤*α*_*k*_≤1 (**Table S1**). If the choosing parent is A^+^, this individual mates preferentially with other A^+^ individuals with probability *α*_1_, and mates randomly with probability 1−*α*_1_, whereas if the choosing parent is A^−^, this individual mates preferentially with other A^−^ individuals with probability *α*_2_, and mates randomly with probability 1−*α*_2_. There are sixteen possible mating pairs from the four phenotypes described, and we use the notation *m*_*i,j*_ to indicate the frequency of a mating between a choosing parent of type *i* and the second parent of type *j* where *i, j* = {1, 2, 3, 4} (**Table S1**); for example, *m*_1,3_ represents the mating frequency of V^+^A^+^ (*x*_1_) and V^−^A^+^ (*x*_3_).

Since the two traits (**A** and **V**) are transmitted vertically, for each phenotype we must specify the probability that the mating produces an offspring of that phenotype. The vaccine confidence trait (A^+^) is transmitted with probability *C*_*n*_, and the vaccine hesitancy trait (A^−^) is transmitted with probability 1*−C*_*n*_ (for *n* = {0, 1, 2, 3} as shown in **Tables 2** and **Table S2**). If *C*_0_ = 0, two A^−^ parents will always produce A^−^ offspring, and if *C*_3_ = 1, two A^+^ parents will always produce A^+^ offspring. However, if *C*_0_ > 0, two A^−^ parents can produce A^+^ offspring at some probability, and similarly if *C*_3_ < 1, two A^+^ parents can produce A^−^ offspring with some probability.

Transmission of vaccination (V^+^ with probability *B*_*m,n*_ for *m, n* = {0, 1, 2, 3}; **Table 1**) is more complex, since parents’ vaccine attitudes (**A**), in addition to their own vaccination states (**V**), can influence their behavior in vaccinating their offspring via a set of “influence parameters” that inform vaccination probabilities. The probability that each mating pair produces an offspring with the V^+^ trait (i.e. vaccinates their offspring) is a scaled product of the influence of parental attitudes (*c*_*n*_ for *n* = {0, 1, 2, 3}) and the influence of parental vaccination states (*b*_*m*_ for *m* = {0, 1, 2, 3}) (**Tables 2 and Table S2**). For example, for mating pair V^+^A^+^ × V^+^A^*−*^, the combined vaccination states (V^+^ × V^+^) will influence vaccination behavior by *b*_3_, and the combined attitude states, (A^+^ × A^*−*^), will influence vaccination behavior by *c*_2_. Therefore, a V^+^A^+^ × V^+^A^*−*^ mating will produce a V^+^ offspring with probability 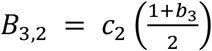; this pair will also produce an A^+^ offspring with probability *C*_2_ based on their combined attitude states. Thus, according to the model, this pairing will produce a V^+^A^+^ offspring with probability *B*_3,2_*C*_2_ and a V^+^A^*−*^ offspring with probability *B*_3,2_(1−*C*_2_). We note that assortative mating (*α*_*k*_>0) will increase the frequency of matings between individuals that share an attitude trait, with these non-random interactions in turn skewing vaccination outcomes toward those of same-state couples (via *c*_0_ and *c*_3_).

**Table 1:** List of parameters, their definitions, and default or initial values.

| Parameter | Meaning | Parameter | Meaning |
| --- | --- | --- | --- |
| **V** | Vaccination state ($V^+$ vaccinated, $V^-$ unvaccinated) | **A** | Vaccine attitude ($A^+$ confident, $A^-$ hesitant) |
| $m_{ij}$ | Mating frequencies (given in **Table S1**) | $\alpha_k$ | Assortative mating parameter (homophily)<br>**Default:** $\alpha_1 = 0$, $\alpha_2 = 0$ |
| $B_{m,n}$ | Probability that parental pairs vaccinate their children, which depends upon the parents' vaccination states ($b_m$) and vaccine attitude ($c_n$) (given in **Table S2**) | $C_n$ | Probability that parental pairs transmit vaccine confidence to their children<br>**Default:** $C_0 = 0.01$, $C_1 = C_2 = 0.5$, $C_3 = 0.99$ |
| $b_m$ | Probability that parental pairs support offspring vaccination given their vaccination states<br>**Default:** $b_0 = 0.01$, $b_1 = b_2 = 0.5$, $b_3 = 0.99$ | $c_n$ | Probability that parental pairs support offspring vaccination given their vaccine attitude<br>**Default:** $c_0 = 0.01$, $c_1 = c_2 = 0.5$, $c_3 = 0.99$ |
| $\sigma$ | Comprehensive selection coefficient for $V^+$, dependent on the population-wide vaccination rate (see **Figure 1**) | $\sigma_{\max}$ | The highest additional benefit that can be conferred by vaccination<br>**Default:** $\sigma_{\max} = 0.1$ |
| **Initial Phenotype Frequencies** | | $x_1(V^+A^+) = 0.81$, $x_2(V^+A^-) = 0.1$, $x_3(V^-A^+) = 0.07$, $x_4(V^-A^-) = 0.02$ | |

**Table 2:** Presence (+) and absence (–) subscript assignments. Demonstrating the trait presence (+) and absence (–) combinations associated with m, n subscripts. For example, the + × – combinations is associated with m and n subscript value 2: an A^+^ × A^−^ pairing transmits A^+^ at probability *C*_2_. This rule applies to parameters *C*_*n*_, *b*_*m*_, *B*_*m,n*_, *c*_*n*_, as shown in **Table S2**.

| Subscript Value ($m, n$; e.g. $b_m, C_n$) | Associated Pairing (e.g. $V \times V, A \times A$) |
| --- | --- |
| 0 | $- \times -$ |
| 1 | $- \times +$ |
| 2 | $+ \times -$ |
| 3 | $+ \times +$ |

Transmission and influence probabilities are constant throughout a single simulation, with values ranging from 0 to 1. At default settings, the influence parameters *b*_*m*_ and *c*_*n*_, and the transmission parameter *C*_*n*_ would take the following values: *C*_0_, *b*_0_, *c*_0_ = 0.01; *C*_1_, *C*_2_, *b*_1_, *b*_2_, *c*_1_, *c*_2_ = 0.5; and *C*_3_, *b*_*3*_, *c*_*3*_ = 0.99. In our model, the influence of parental vaccine beliefs (*c*_*n*_) is greater than the influence of their own vaccination status (*b*_*n*_) on their likelihood of vaccinating their offspring, so offspring vaccination is guaranteed at some probability only if *c*_*n*_ > 0.

Transmission and influence probabilities are constant throughout a single simulation, with values ranging from 0 to 1. At default settings, the influence parameters *b*_*m*_ and *c*_*n*_, and the transmission parameter *C*_*n*_ would take the following values: *C*_0_, *b*_0_, *c*_0_ = 0.01; *C*_1_, *C*_2_, *b*_1_, *b*_2_, *c*_1_, *c*_2_ = 0.5; and *C*_3_, *b*_*3*_, *c*_*3*_ = 0.99. In our model, the influence of parental vaccine beliefs (*c*_*n*_) is greater than the influence of their own vaccination status (*b*_*n*_) on their likelihood of vaccinating their offspring, so offspring vaccination is guaranteed at some probability only if *c*_*n*_ > 0.

The cultural selection pressure on vaccination is given by the parameter σ, such that the frequency of the V^+^A^+^ and V^+^A^−^ phenotypes are multiplied by 1+σ after vertical cultural transmission has occurred. At the end of each timestep, the frequency of each phenotype is divided by the sum of all four frequencies, ensuring that the frequencies sum to 1. This cultural selection coefficient is implemented in the same way as a selection coefficient in a population-genetic model, but unlike the latter, it is structured to encompass both biological fitness and cultural selection pressures, including perceived risks or benefits of the vaccine itself, personal cost-benefit analyses of preventative health behaviors, and the structural or societal-level factors influencing vaccination rates [32,33]. This parameter modulates whether there are more or fewer vaccinated individuals than expected: in other words, when σ>0, vaccinated individuals are more common in a set of offspring than would be expected strictly based on the beliefs and vaccination statuses of their parents. We calculate σ in each timestep as a function of the current vaccination coverage (frequency of V^+^, i.e. *x*_1_ + *x*_2_), and in each simulation we specify σ_*max*_ as the maximum cultural selection pressure of getting vaccinated (−1≤*σ*_*max*_≤1) (see the cultural selection coefficient function in **Figure 1**). This function was constructed by fitting a curve to pre-specified conditions that incorporate assumptions from evolutionary game theory (e.g. that herd immunity decreases the incentive to vaccinate [34]): we assume that when vaccination coverage is low, the real and perceived benefits of vaccination are highest, and thus, the cultural selection pressure is near σ_max_, however, as vaccination coverage increases, the perceived benefits of vaccination decrease and the cultural selection pressure is reduced (**Figure 1**).

**Figure 1:**
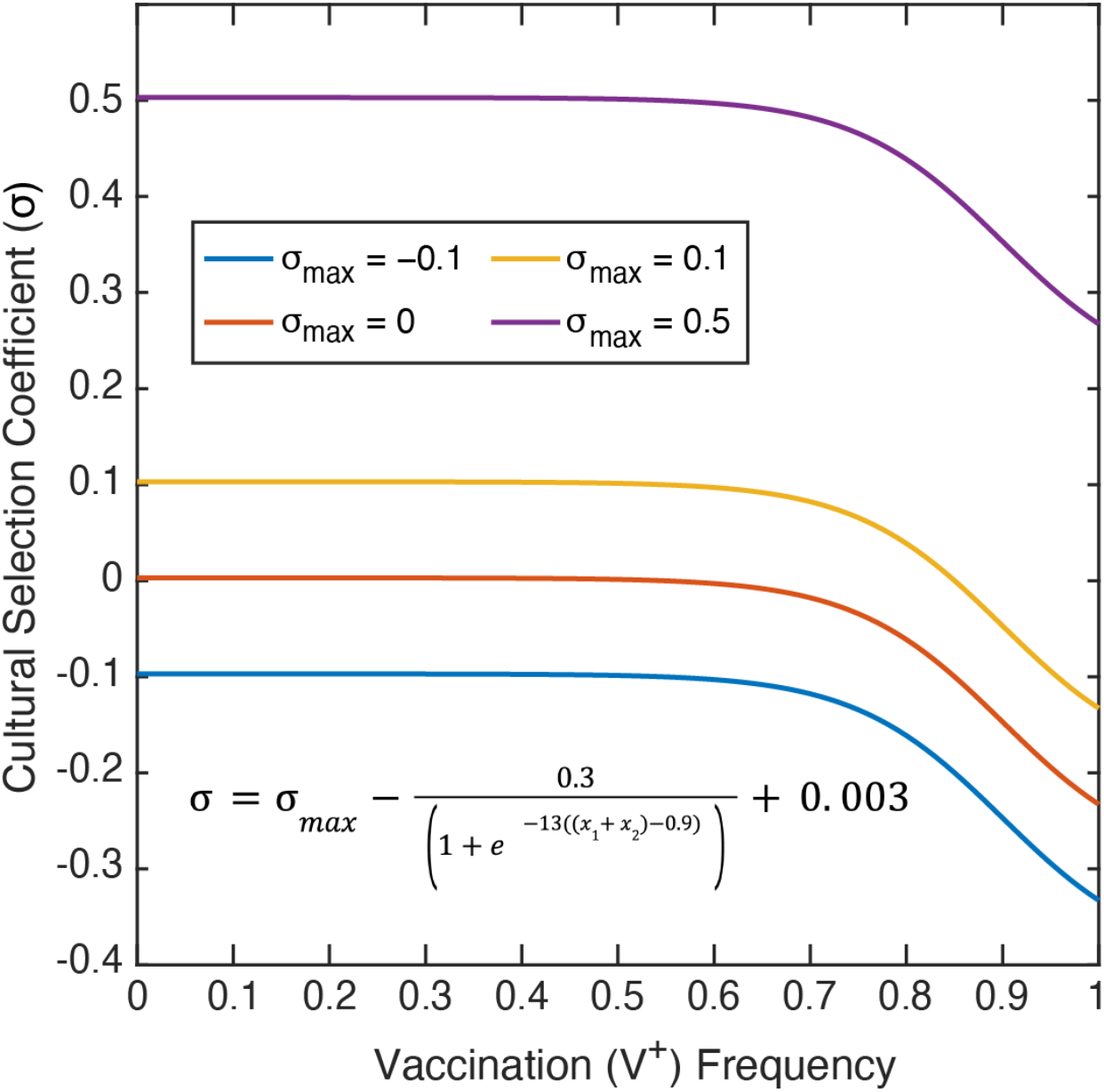
Cultural selection coefficient function. The cultural selection coefficient function was constructed by fitting a curve to specified conditions, and considers both health and non-health related effects. The selection coefficient (σ; vertical axis) is dependent on the frequency of vaccinated individuals (V^+^) in the population (horizontal axis). σ_max_ is the maximum cultural selection coefficient associated with being vaccinated. Perceived vaccine benefit is reduced as vaccination coverage increases, since the negative effects of the disease will be less apparent. Note: Of the σ_max_ values shown, only σ_max_ = 0.1 allows the cultural selection pressure to be either positive or negative at a given timepoint depending on the frequency of vaccination.

Thus far, we have described vertical cultural transmission from parent to offspring. The model also incorporates a second phase with oblique cultural transmission (i.e. influence from non-parental adults), in which individuals can change their inherited vaccine attitudes (**A**) due to influence from other adults in the population. There are two probabilities associated with attitude modulation: the probability that a vaccine hesitant (A^*−*^) individual adopts the vaccine confident (A^+^) state (A^*−*^ to A^+^ transition probability, given by *A*_*→Confident*_ in **Figure 2**), and the probability that an A^+^ individual adopts the A^*−*^ state (A^+^ to A^*−*^ transition probability, given by *A*_*→Hesitant*_ in **Figure 2**). As with the strength of cultural selection (σ) described previously (**Figure 1**), the probability that offspring change their vaccine attitude is a function of the V^+^ frequency in the population, constructed according to similar assumptions given in **Figure 2**. As the frequency of vaccinated individuals (V^+^) increases in the population, vaccine-confident individuals (A^+^) are more likely to become hesitant (*A*_*→Hesitant*_ probability increases) and vaccine-hesitant individuals (A^−^) are less likely to become confident (*A*_*→Confident*_ probability decreases).

**Figure 2:**
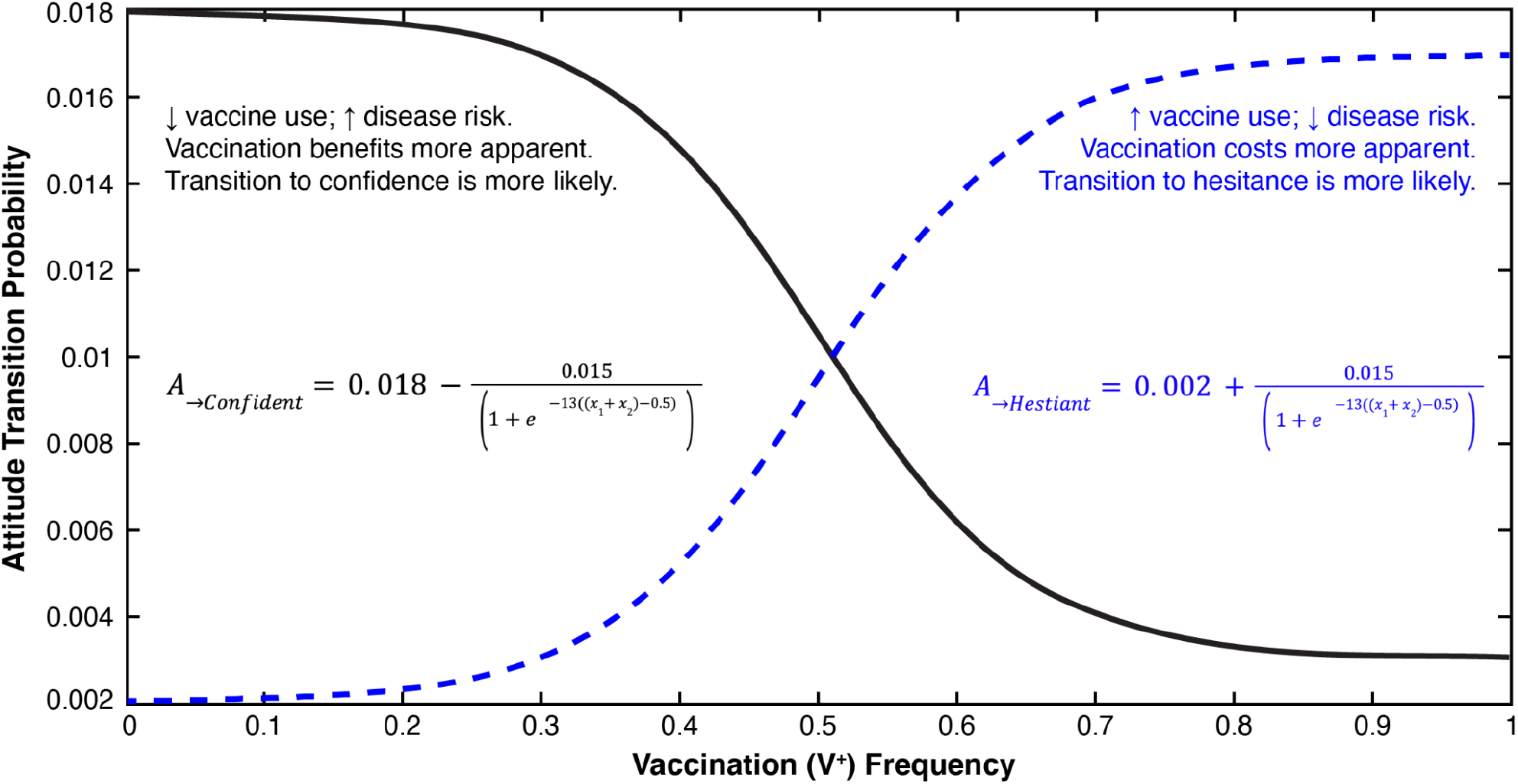
Attitude Transition Probability Function. Attitude transition probability functions were constructed by fitting a curve to specified values based on the assumptions pictured. Attitude transition probability (vertical axis) is a function of the vaccination frequency in the population (V^+^; horizontal axis). The probability that a vaccine hesitant individual adopts vaccine confidence (A^−^ to A^+^ transition probability, shown in black) is determined by the function *A*_*→Confident*_, and the probability that a vaccine confident individual adopts vaccine hesitancy (A^+^ to A^−^ transition probability, shown with a blue dashed line) is determined by the function *A*_*→Hesitant*_.

To compute the frequency of a given phenotype in the next iteration, we sum the probability that each mating pair produces offspring of that phenotype over each of the sixteen possible mating pairs. Cultural selection (*σ*), described above, then operates on offspring with the V^+^ trait. The full recursions, giving *x*_*i*_′ phenotype frequencies in the next iteration in terms of *x*_*i*_ in the current iteration, are given in **Text S1**. If *x*_*i*_′ is equal to *x*_*i*_, the system is at equilibrium. As we do not incorporate a birth-death process or population asynchrony in this model, iterations in the discrete-time format of our model should not be strictly interpreted as years or generations. We instead interpret each iteration broadly as a timeframe in which the specified cultural interactions could occur, which varies among individuals, populations, and cultures. Unless otherwise stated, the model is initialized with phenotypic frequencies based on United States data: *x*_1_ (frequency of V^+^A^+^) = 0.81, *x*_2_ (V^+^A^−^) = 0.1, *x*_3_ (V^−^A^+^) = 0.07, *x*_4_ (V^−^A^−^) = 0.02. These frequencies were estimated using reports of Measles-Mumps-Rubella (MMR) vaccination rates and estimates of vaccine attitude frequencies obtained from various sources in the literature [35,36] and the Centers of Disease Control ChildVax database [37,38].

## 3. Results

To test our model, we first initialized a population with a set of phenotype frequencies and examined the changes in these frequencies over time with a given set of parameters (**3.1**). Then, we evaluated the effects of each parameter by running simulations at multiple parameter combinations and recording the population frequencies of each phenotype once the system approached an equilibrium (**3.2–3.4**). In our first set of simulations (**3.1–3.3**), we include only vertical transmission dynamics, i.e. only parent-to-offspring transmission, varying parameter values in turn to test their effects on population vaccination behavior and attitudes. In the vertical transmission phase of the model, parents choose whether to vaccinate their offspring (i.e., transmit V^+^) or to not vaccinate (V^−^), and parents also transmit a vaccine attitude (confidence, A^+^, or hesitancy, A^−^), each with a specified probability given the phenotypes of the parents. The parental attitude state, vaccination status, assortative mating levels, and cultural selection parameters interact to affect vaccination coverage (frequency of V^+^ in the population) and vaccine confidence (frequency of A^+^).

### 3.1 Temporal dynamics of vaccine-related beliefs and behaviors

To test whether the equilibrium phenotype frequencies were sensitive to starting frequencies, we plotted the dynamics of each phenotype over time at default parameters (given in **Table 1**). For each set of initial phenotype proportions tested, the phenotype frequencies in the population quickly adjusted to approach equilibrium values and then gradually plateaued to a stable equilibrium (vertical transmission: **Figure 3 and Figure S1**, vertical+oblique transmission: **Figure S2**. This demonstrates that equilibrium frequencies of vaccination coverage and vaccine confidence are determined by the parameter conditions rather than by the initial frequencies themselves.

**Figure 3:**
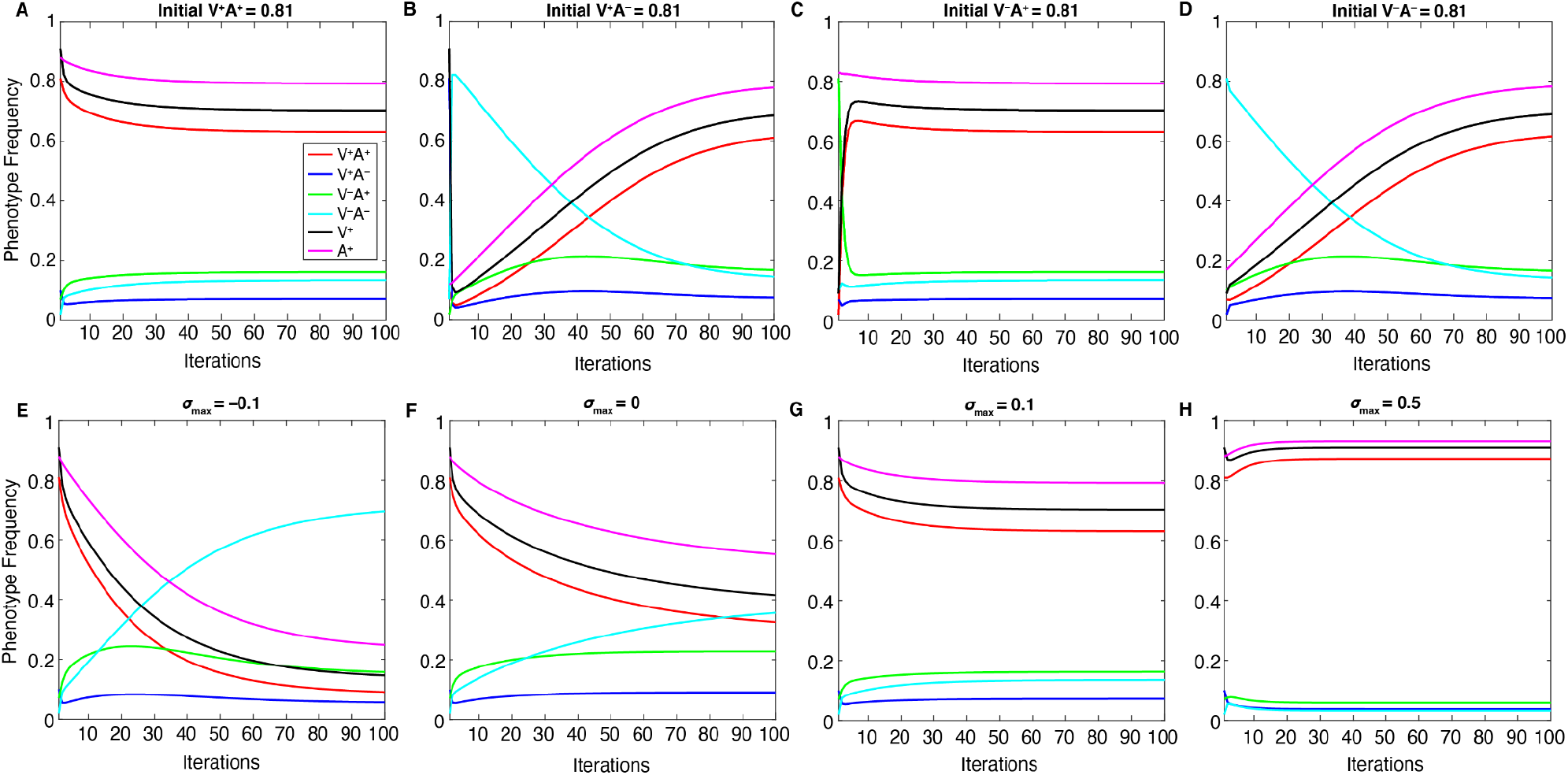
Equilibrium frequencies are determined by the parameter space, not by initial frequencies. The change in each of the four phenotype frequencies and the total V^+^ and A^+^ frequencies (vertical axis) over 100 iterations of the model **with vertical transmission only** (horizontal axis). **Top Row:** Initial frequencies are varied, such that we begin each simulation with a different phenotype at an initial high frequency (0.81): V^+^A^+^ in panel **A**, V^+^A^*−*^ in panel **B**, V^*−*^A^+^ in panel **C**, V^*−*^A^*−*^ in panel **D**; the remaining phenotypes are set to lower frequencies (0.1, 0.07, 0.02). See **Figure S1** for a full listing of these initial frequencies. **Bottom Row:** The maximum cultural selection coefficient (σ_max_) is varied: **E**. σ_max_ = −0.1; **F**. σ_max_ = 0; **G**. σ_max_ = 0.1; **H**. σ_max_ = 0.5. Cultural selection against vaccinated individuals increases the frequency of V^*−*^A^*−*^, decreasing the other frequencies (**E**), whereas increased cultural selection favoring vaccinated individuals increases V^+^A^+^ frequencies while decreasing the other frequencies (**F, G, H**). In all panels, the remaining parameters are held at default values (**Table 1**).

When two parameters in particular are varied—maximum cultural selection (σ_max_) or confidence transmission (*C*_1_ = *C*_2_)—we observe a trade-off between the V^−^A^−^ phenotype, which dominates at lower values of these parameters, and the V^+^A^+^ phenotype, which dominates at higher values (**Figures 3-4**). Interestingly, the “conflicting” phenotypes (when an individual’s attitude toward vaccinating their children does not match their own vaccination state: V^−^A+ and V^+^A^−^) are present at their highest frequencies at neutral cultural selection (σ_max_ = 0, **Figure 3F**) and/or neutral confidence transmission (*C*_1_ = *C*_2_ = 0.5, **Figure 4B**). Vaccinated individuals have the same fitness regardless of their attitude (i.e. V^+^A^+^ bears the same selection pressure as V^+^A^−^), so it is worth noting that at higher levels of confidence transmission and cultural selection, V^+^A^+^ increases in frequency but V^**+**^A^−^ decreases in frequency (compare **Figure 3F-G, Figure 4B-C**). This pattern seems to reflect their differing likelihoods of vaccinating their offspring: across all possible partners, vaccinated but vaccine-hesitant parents (V^+^A^−^) are less likely to vaccinate their offspring than vaccinated and vaccine-confident parents (V^+^A^+^), resulting in more V^−^ offspring.

**Figure 4:**
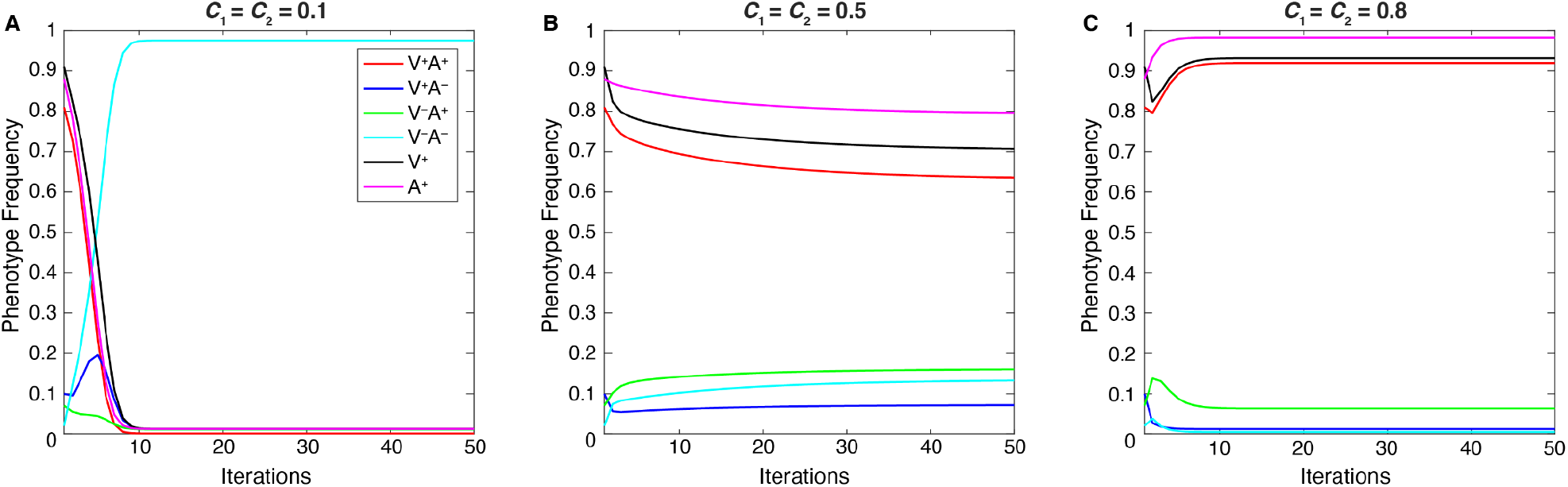
Temporal Effects of Confidence Transmission. The change in phenotype frequencies over 50 iterations as vaccine confidence transmission in mixed-attitude couples (*C*_1_ = *C*_2_) is varied (**A**. *C*_1_ = *C*_2_ = 0.1; **B**. *C*_1_ = *C*_2_ = 0.5; **C**. *C*_1_ = *C*_2_ = 0.8) **with vertical transmission only**, while other parameters are held at default values (**Table 1**). The population equilibrates at over 90% A^−^V^−^ at low confidence transmission (A). Increasing the probability of confidence transmission results in less vaccine hesitancy and, in turn, higher vaccination frequencies (V^+^A^+^).

Thus, when V^+^ is favored by cultural selection, there is indirect selection against the vaccinated but vaccine-hesitant (V^+^A^−^) phenotype (**Figure 3E-H**). Similarly, indirect selection against V^−^A^+^ occurs when V^−^ is favored by cultural selection (**Figure 3E**): compared to **Figure 3F**, we observe an increase in V^−^A^−^ individuals but a decrease in V^−^A^+^ individuals, who because of their vaccine confidence have more V^+^ offspring, which are culturally disfavored in this environment. When cultural transmission from non-parental adults (oblique transmission) was included, described in following sections, we observed similar patterns, but the final equilibria were more likely to be polymorphic, with vaccinated, unvaccinated, confident, and hesitant phenotypes stabilizing at more moderate frequencies than they would have with only vertical transmission (Compare **Figures 3-4 to Figures S3-S4**).

Low confidence transmission (*C*_1_ = *C*_2_ = 0.1, **Figure 4A**) increases the frequency of vaccine hesitancy (A^−^) in the population over time, increasing the probability that more couples choose not to vaccinate their offspring. However, the increase in vaccine hesitancy does not occur equally in vaccinated and unvaccinated individuals: A^−^ frequency may increase overall in this environment, but V^+^A^−^ frequencies are lower and V^−^A^−^ frequencies are higher (compared to **Figure 4B-C** and **Figure S4**). At neutral confidence transmission probabilities (i.e. when couples with one confident and one hesitant parent are equally likely to transmit either attitude), there is a higher chance that the vaccinated but vaccine-hesitant (V^+^A^−^) phenotype is replenished. However, if vaccine confidence is highly transmitted (*C*_1_ = *C*_2_ = 0.8), the V^+^A^−^ frequency will be reduced, as this phenotype is more likely to produce A^+^ offspring than A^−^, thus increasing V^+^A^+^ phenotype frequencies in the population (**Figure 4** and **Figure S4**). If we turn to the other conflicting phenotype, unvaccinated but vaccine-confident (V^−^A^+^) individuals become more common when A^+^ increases in frequency in the population as *C*_1_ = *C*_2_ increases from 0.1 to 0.5 (**Figure 4** and **Figure S4**). In contrast, higher vaccine confidence transmission (*C*_1_ = *C*_2_ = 0.8) can lead to a vaccination-promoting environment in which V^−^ frequencies are reduced over time; thus the V^−^A^+^ phenotype becomes rare and V^+^A^+^ predominates (**Figure 4** and **Figure S4**).

### 3.2 Parent-to-Offspring Interactions (Simulations with vertical transmission only)

Since our assessment of the temporal dynamics (**3.1**)demonstrated that our simulations approach stable equilibria, we then modulated different sets of parameters and recorded the phenotype frequencies at equilibrium, generating heat maps showing the results across a range of parameters. In the first of these, we tested the relationship between vaccination probability and vaccine confidence transmission. To directly alter vaccination probabilities while still accounting for the couple’s vaccine attitudes, we set ranges of values for *B*_*m,n*_ that vary along the horizontal axis of **Figure 5**, with the vaccination probability for two hesitant parents (e.g. *B*_0,0_) on the lower end of the range and the vaccination probability for two confident parents (e.g. *B*_3,3_) on the higher end of the range (**Table S3**). Confidence transmission probabilities are also structured in this “range shift” manner (**Figure 5A-B, Table S3**). If we vary both confidence transmission parameters and vaccination probability parameters by implementing range shifts in both *C*_*n*_ and *B*_*m,n*_, we observe a positive interaction between confidence transmission and vaccination probability: vaccination coverage increases as either of these parameters are increased (**Figure 5A**).

**Figure 5:**
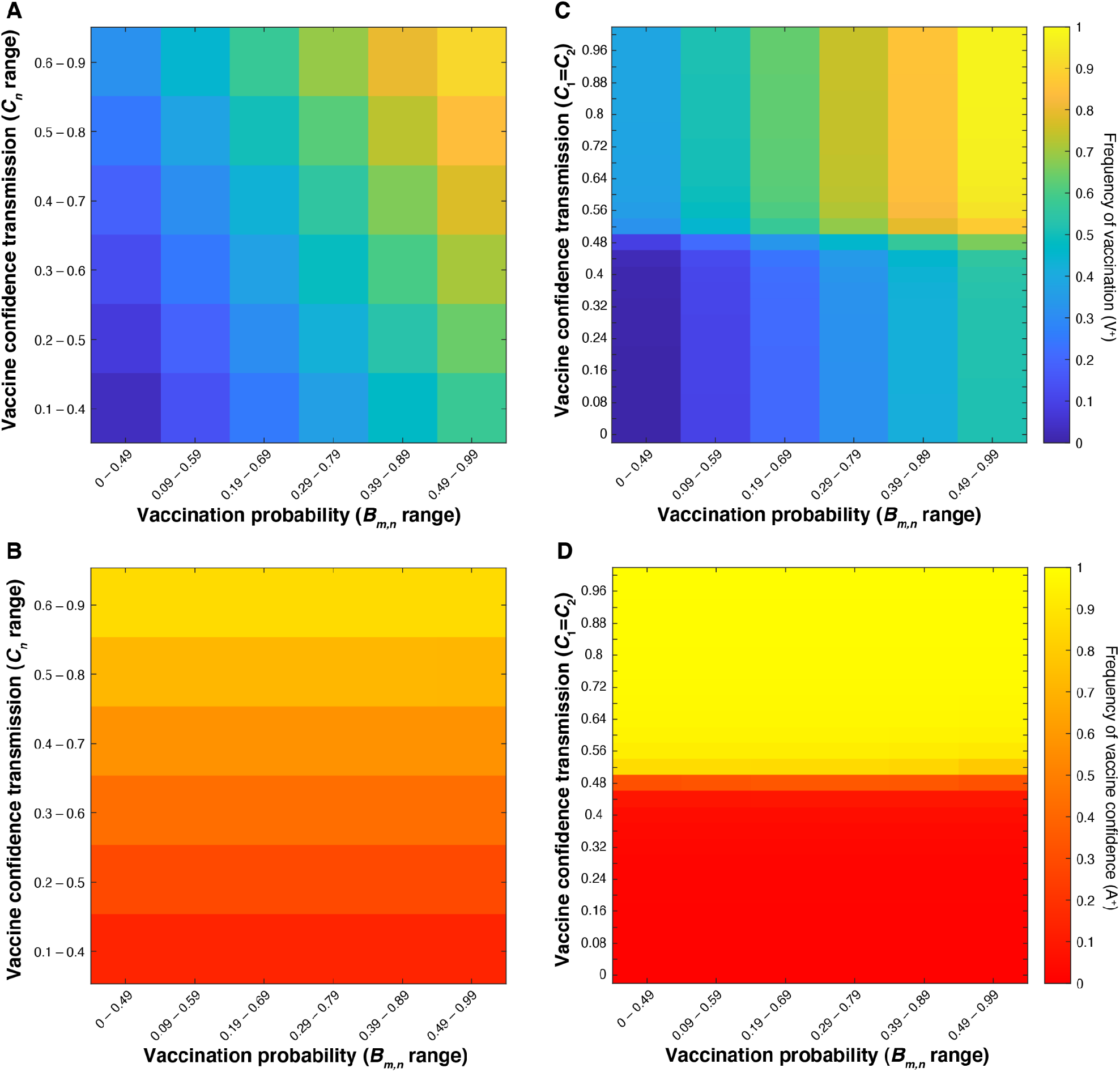
Vaccination coverage levels are determined by an interaction between confidence transmission and vaccination probability. Heatmaps showing final vaccination coverage (**A, C**) and corresponding vaccine confidence (**B, D**) after 100 time-steps with **no oblique transmission**. In **A** and **B**, confidence transmission probabilities (*C*_*n*_) are set within the range indicated on the vertical axis, and vaccination probabilities (*B*_m,n_) are set within the range indicated on the horizontal axis with *B*_0,0_, *B*_1,0_, *B*_2,0_ and *B*_3,0_ taking the lowest value and *B*_3,3_ taking the highest value (**Table S3**). In **C** and **D**, confidence transmission in mixed-attitude couples (*C*_1_ = *C*_2_) is varied along the vertical axis, while the vaccination probabilities (*B*_*m,n*_) are set within the range indicated on the horizontal axis as in **A** and **B**. (**Table S3**). We show increased equilibrium vaccination coverage with increasing vaccination probability and confidence transmission probability ranges, while confidence levels are primarily dictated by proportion of the population transmitting confidence or hesitancy.

However, couples with mixed vaccination and/or attitude states (V^+^ × V^−^, A^+^ × A^−^) are assumed to be more variable in their decision to vaccinate their offspring than parents who share the same state. Thus, in the simulations that follow, we primarily modulated the specific probabilities associated with these mixed-state pairings. In **Figure 5C-D**, we varied vaccination probabilities (*B*_*m,n*_) across the full range of individuals but modulated confidence transmission probabilities only for mixed-attitude couples (*C*_1_ = *C*_2_), i.e. those with one vaccine-hesitant parent and one vaccine-confident parent. In these tests, we observe increasing equilibrium vaccination coverage as *B*_*m,n*_ probabilities increase, with higher coverage in high-confidence transmission environments (**Figure 5C-D**).

In both aforementioned simulations (**Figure 5**), we confirm vaccination coverage levels are determined by an interaction between confidence transmission and vaccination probability, whereas confidence levels are dictated primarily by levels of confidence transmission. In sum, the degree to which parents with mixed vaccine-hesitant and vaccine-confident attitudes transmit vaccine confidence instead of vaccine hesitancy to their offspring is a key factor in determining population trait majorities which can drastically shift population dynamics.

We compared the effects of varying the confidence transmission probabilities for mixed-attitude couples (*C*_1_ and *C*_2_) in combination with multiple factors: 1) the maximum cultural selection coefficient (σ_max_) (**Figure 6A-B**), 2) the vaccination influence parameters *b*_1_ and *b*_2_ (**Figure 6C-D**), 3) the attitude influence parameters *c*_1_ and *c*_2_ (**Figure 6E-F**), and 4) the vaccination probabilities of couples with mixed states, *B*_1,1_, *B*_1,2_, *B*_2,1_, *B*_2,2_ (**Figure 6G-H**). In each examination, we observed a *C*_n_ threshold: there is a mid-range value of *C*_*n*_ at which vaccination coverage and vaccine confidence traits are polymorphic (i.e. both forms of each trait coexist in the population), separating definitive high (⪆80%) and low (⪅30%) levels of vaccination coverage and confidence. This *C*_*n*_ threshold value is more sensitive to σ_max_ than to *b*_*m*_, *c*_*n*_, or *B*_*m,n*_: the threshold value is lowered as σ_max_ increases (diagonal line in **Figure 6A-B**). Although vaccination probability (*B*_*m,n*_) is dependent on both *c*_*n*_, the influence of parental vaccine attitude, and *b*_*m*_, the influence of parental vaccination state (**Table S2**), modulating either type of influence of mixed-state parents has little effect on the level of vaccination coverage and negligible effects on confidence levels at each non-threshold *C*_*n*_ (**Figure 6C-F**).

**Figure 6:**
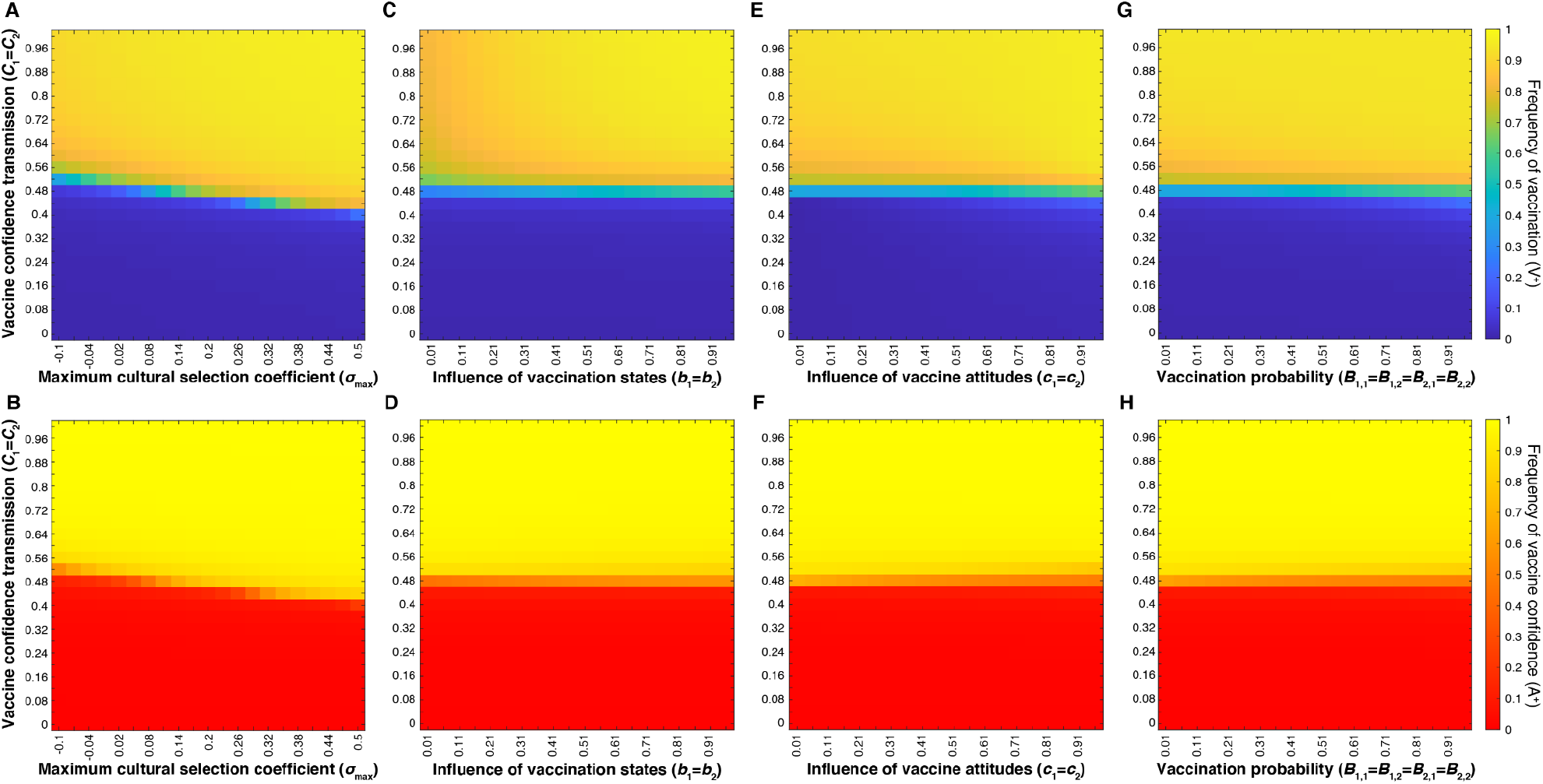
Vaccine Confidence Transmission Dictates Vaccination Coverage and Confidence Levels. Heatmaps showing final vaccination coverage and vaccine confidence after 100 time-steps with **no oblique transmission**, only parent-to-offspring transmission. The top row (**A, C, E, G**) shows vaccination coverage (i.e. frequency of V^+^ in the population) with low coverage in blue and high coverage in yellow; the bottom row (**B, D, F, H**) shows the corresponding final vaccine confidence (i.e. frequency of A^+^), with low confidence in red and high confidence in yellow. Unless varied on the horizontal or vertical axis, other parameters are set to the default values given in **Table 1**. In our model, parents’ likelihood of vaccinating their children depends on both their vaccination state and their attitude state. This figure shows that the strength of parental transmission of vaccine confidence (*C*_*n*_) has a much stronger effect on the equilibrium levels of both vaccine coverage (V^+^) and confidence (A^+^) than other parameters: the maximum cultural selection coefficient, σ_max_ (**A**,**B**), the influence of parental vaccination state, *b*_*m*_ (**C, D**), the level of influence of parental vaccine attitudes on their vaccination behaviors, *c*_*n*_ (**E**,**F**), and the probability that mixed-state parents vaccinate their offspring *B*_*m,n*_ (**G**,**H**).

Interestingly, direct modulation of the mixed-state couple vaccination probability (*B*_1,1_ = *B*_1,2_ = *B*_2,1_ = *B*_2,2_) also has little power in affecting coverage and confidence levels at equilibrium (**Figure 6G-H**). We hypothesize that predominantly high or predominantly low confidence transmission within a population reduces the occurrence of “mixed-state” pairings, i.e. if the majority of the population becomes confident or hesitant, there are fewer confident-hesitant and vaccinated-unvaccinated pairings. Thus, the effect of modulating mixed-state vaccination probabilities (*B*_1,1_, *B*_1,2_, *B*_2,1_, *B*_2,2_) is significantly minimized as these couples approach low frequencies in the population, and confidence transmission dominates the vaccination patterns.

Next, we hold vaccine confidence transmission (*C*_*n*_) at default probabilities, reminiscent of Mendelian transmission, such that two vaccine confident or two vaccine hesitant parents predictably transmit their vaccine attitude, and parents with differing vaccine attitudes each have a ∼50% chance of transmitting their own state, e.g. *C*_0_ near 0, *C*_1_ and *C*_2_ at 0.5, *C*_3_ near 1 **(Table 1)**.

We then varied cultural selection in combination with vaccination-associated probabilities (*b*_*m*_, *c*_*n*_, *B*_*m,n*_). With *C*_*n*_ held constant, cultural selection (σ_max_) is the primary factor determining vaccination coverage and confidence levels (**Figure 7**). Raising the maximum cultural selection coefficient increases the equilibrium level of vaccination coverage and vaccine confidence across various levels of vaccination state influence (*b*_*m*_) (**Figure 7A-B**), vaccination attitude influence (*c*_*n*_) (**Figure 7C-D**), and vaccination probability (*B*_*m,n*_) (**Figure 7E-F**). Unlike in **Figure 6**, vaccine confidence does not always mirror vaccination coverage across all levels of attitude influence (*c*_*n*_) or vaccination probabilities. Instead, vaccine confidence levels decline with increased *c*_*n*_ and increased *B*_*m,n*_ for σ_max_ ⪅ 0.3 (**Figure 7D, F**), as well as for both increased *c*_*n*_ and increased *b*_*m*_ (**Figure S5**). This dynamic is interesting as these parameters inform vaccination behavior, hinting that high vaccination rates could reduce a populations’ expected vaccine confidence. Vaccination coverage and vaccine confidence remain low when cultural selection does not favor vaccination (σ_max_ ⪅ 0), i.e. parents vaccinate their children at or below the levels expected based on cultural transmission rates.

**Figure 7:**
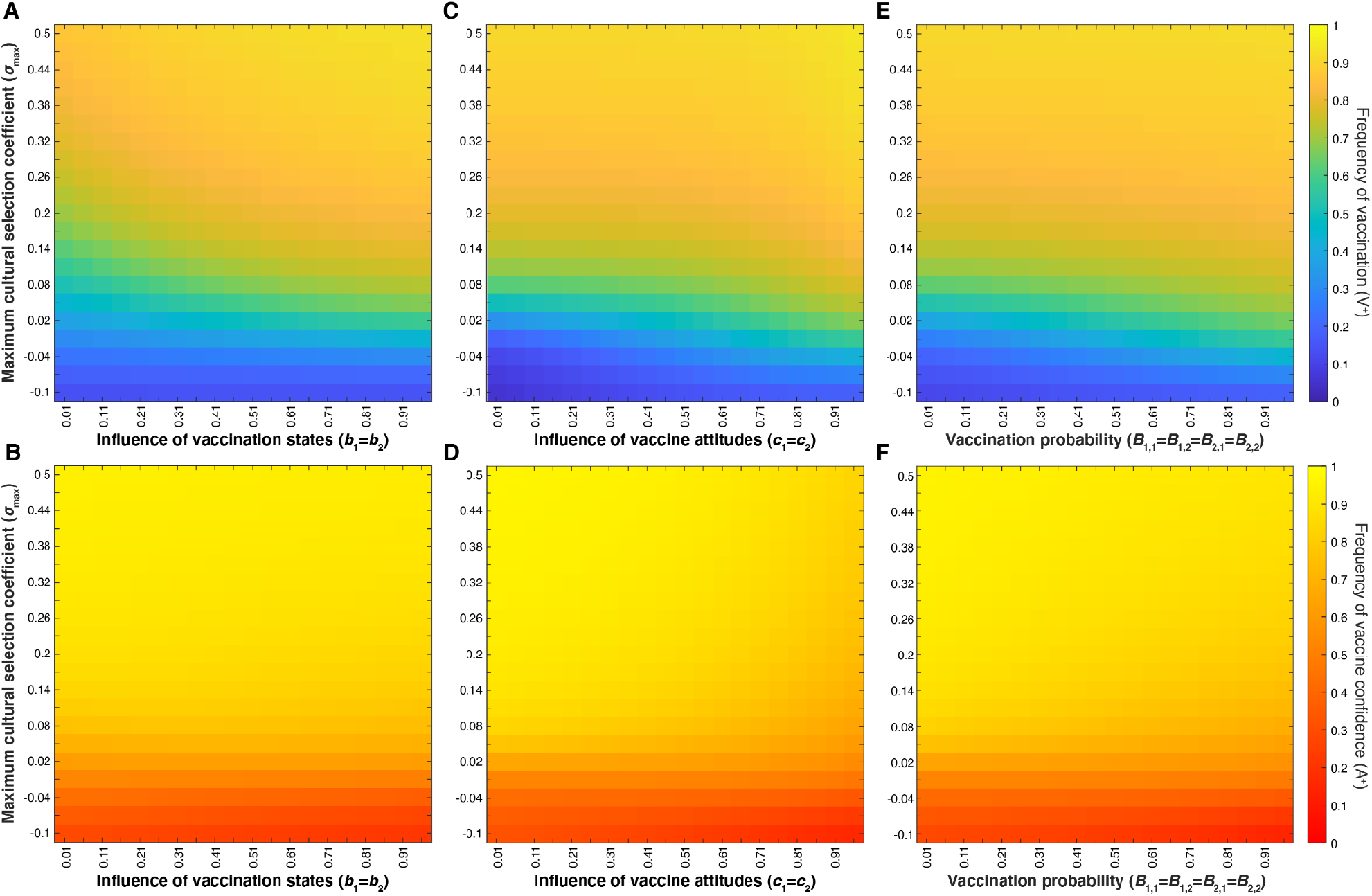
Cultural Selection Influences Vaccination Coverage and Vaccine Confidence. Heatmaps showing final vaccination coverage (**A, C, E**) and final vaccination confidence (**B, D, F**) after 100 time-steps with **no oblique transmission**, only parent-to-offspring transmission. As in previous figures, parameters not varied here are given in **Table 1**. Parents’ likelihood of vaccinating their children depends on both their vaccination state and their attitude state. At default probabilities of vaccine confidence transmission (*C*_n_ values in **Table 1**), these figures show that modulating the maximum cultural selection coefficient affects the equilibrium levels of vaccination coverage and vaccine confidence across the range of specified parameters: parental vaccination state influence, *b*_*m*_ (**A, B**), parental attitude state influence, *c*_*n*_ (**C**,**D**), and offspring vaccination probability, *B*_*m,n*_ (**E**,**F**). Unless directly modulated (as in panels **E-F**), *B*_*m,n*_ varies with *b*_*m*_ and 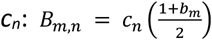.

### 3.3 Offspring can Change their Inherited Hesitancy State (Vertical and Oblique Dynamics)

Increased exposure to the attitudes of the broader community (i.e. oblique cultural transmission from non-parental adults in the population) could influence and change vaccination beliefs inherited in childhood. Therefore, we next included these oblique effects in our model to understand how they might modulate vaccine confidence and vaccination coverage levels. In the oblique transmission phase of the model, offspring can change their vaccine attitude with some probability based on the frequency of vaccination in the population (**Figure 2**). Thus, in addition to the vertical transmission of attitudes and behaviors, phenotype frequencies are further affected by the probability that adult offspring change their attitude (i.e. transition from vaccine confident (A^+^) to hesitant (A^−^) and vice versa). By modulating the attitude transition probabilities according to the vaccination coverage, we assume that when vaccine coverage (V^+^ frequency, *x*_1_ + *x*_2_) is low, disease occurrence is high and the negative effects of the disease are experienced widely, thus the benefits of being vaccinated (and the costs of not being vaccinated) are more evident [39,40]. As vaccination coverage (V^+^) increases in the population, and thus disease occurrence is low, the benefits to being vaccinated are less obvious, while low-probability costs such as adverse reactions become more apparent and could be perceived as being riskier than the disease itself.

The addition of oblique dynamics produces a pattern of vaccination coverage and vaccine confidence similar to that of simulations run with solely vertical transmission (**Figure 6 and Figure 7 compared to Figure 8 and Figure 9, and Figures 3-4** compared to **Figures S2-4**)—the level of (vertical) vaccine confidence transmission still largely determines the level of vaccination coverage and vaccine confidence (**Figure 8**). However, oblique cultural influences expanded the polymorphic space, resulting in a wider range of intermediate *C*_*n*_ in which the different states of each trait (vaccinated, unvaccinated, confident, and hesitant) are present in the population in roughly equal proportions. In other words, there is a wider horizontal stripe of moderate values between the definitively high and definitively low equilibrium frequencies in **Figure 8** than in **Figure 6**). The addition of oblique transmission appears to lead to less polarized results overall, moving the equilibrium levels of vaccination coverage and vaccine confidence away from extreme values.

**Figure 8:**
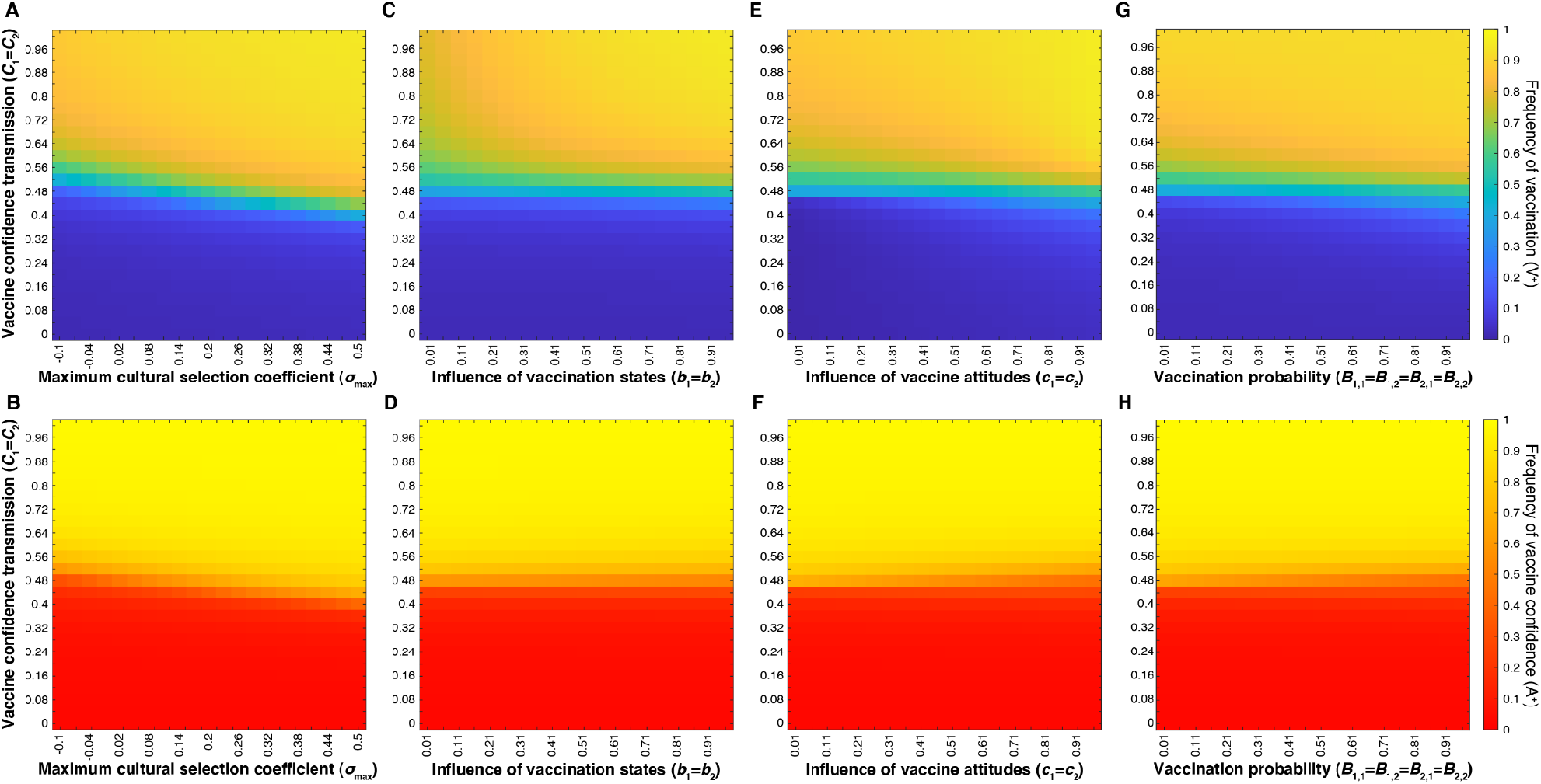
Vaccine confidence transmission dictates vaccination coverage and confidence levels (with oblique transmission). Heatmaps showing final vaccination coverage (i.e. frequency of V^+^ in the population, with low coverage in blue and high coverage in yellow (**A, C, E, G**)) and final vaccine confidence (i.e. frequency of A^+^, with low confidence in red and high confidence in yellow (**B, D, F, H**)) after 100 time-steps in which oblique transmission of vaccine attitude can occur after parent-to-offspring transmission. The likelihood that individuals change their vaccine beliefs depends on the current vaccination coverage of the population (**Figure 2**). Unless varied on the horizontal or vertical axes, other parameters are set to the default values given in **Table 1**. Parents’ likelihood of vaccinating their children depends on both their vaccination state and their attitude state; these figures show that the strength of parental transmission of vaccine confidence (*C*_*n*_) has a much stronger effect on the equilibrium levels of both vaccine coverage (V^+^) and confidence (A^+^) than other tested parameters do: the maximum cultural selection coefficient, σ_max_, (**A**,**B**), the influence of parental vaccination state, *b*_*m*_, (**C, D**), the level of influence of parental attitudes on their vaccination behaviors, *c*_*n*_, (**E**,**F**), and offspring vaccination probability, *B*_*m,n*_ (**G**,**H**).

**Figure 9:**
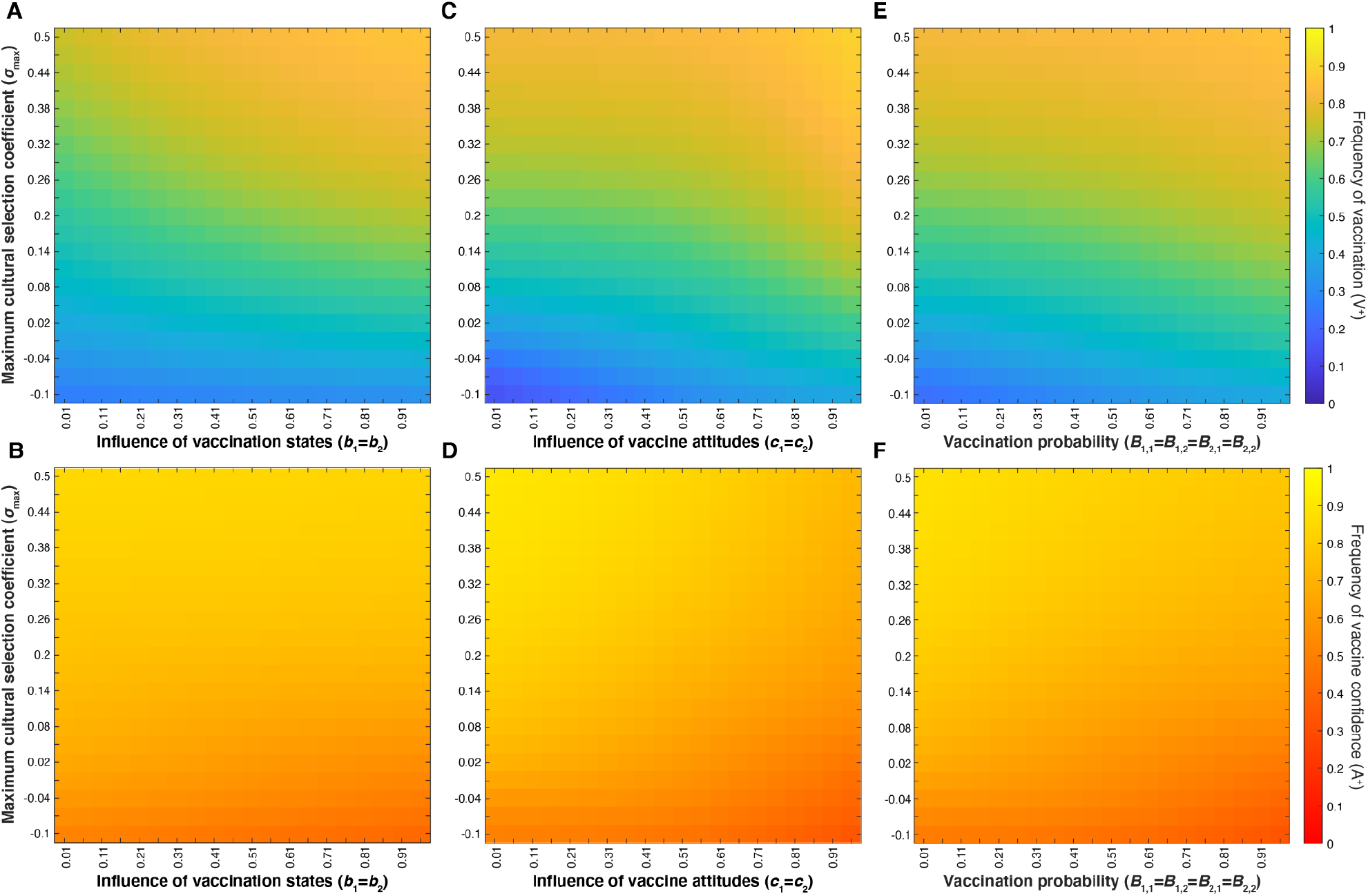
Cultural selection influences vaccination coverage and vaccine confidence (with oblique transmission). Heatmaps showing final vaccination coverage (**A, C, E**) and final vaccination confidence (**B, D, F**) after 100 time-steps **with oblique transmission**. As in previous figures, parameters not varied are given in **Table 1**. Parents’ likelihood of vaccinating their children depends on both their vaccination state and their attitude state. At default probabilities of vaccine confidence transmission (*C*_*n*_), these figures show that modulating the maximum cultural selection coefficient affects the equilibrium levels of vaccination coverage and vaccine confidence across the range of specified parameters: parental vaccination state influence, *b*_*m*_ (**A, B**), parental attitude state influence, *c*_*n*_ (**C**,**D**), and offspring vaccination probability, *B*_*m,n*_ (**E**,**F**). Unless directly varied (as in panels **E-F**), *B*_*m,n*_ varies as *b*_*m*_ and *c*_*n*_ are varied, as shown in **Table 1**.

With neutral confidence transmission (*C*_1_ = *C*_2_ = 0.5), we also observe an expansion of the polymorphic space when we modulate cultural selection (σ_max_) alongside the influence and transmission parameters (**Figure 9**). Interestingly, in the cultural environment defined by this parameter space, we observe a pattern that deviates from the expected association between high vaccine confidence and high vaccination coverage: as the influence of vaccine attitudes (*c*_*n*_) and vaccination probabilities (*B*_*m,n*_) increase (**Figure 9C-F**), the population’s equilibrium vaccination coverage increases while its vaccine confidence decreases. This pattern persisted across all tested levels of maximum cultural selection (σ_max_) (**Figure 9C-F**). In other words, we observe higher levels of confidence when the influence of vaccine attitude is low (**Figure 9D**) and vaccination probabilities are low (**Figure 9F**) than we do at higher values.

We explored the interaction between the influence parameters, *b*_*m*_ and *c*_*n*_, at various maximum cultural selection coefficients (σ_max_) (**Figure 10**). Vaccination coverage and vaccine confidence equilibrate at mid-range frequencies (between 0.3 and 0.8) across the range of *b*_*m*_ and *c*_*n*_, indicating that these trait frequencies are not particularly sensitive to either parameter. Cultural selection favoring vaccination increases the equilibrium level of vaccination coverage and vaccine confidence (**Figure 10 and Figure S6**). The most notable deviation between equilibrium confidence and vaccination frequencies occurs at the intersection of the highest influence parameter values (*b*_*m*_ and *c*_*n*_), circumstances in which the parents’ vaccination states and vaccine attitudes overwhelmingly support offspring vaccination. In this top right section of the heat maps, vaccination coverage is high while vaccine confidence is lower, indicating a behavioral pattern in which mixed-trait couples are more inclined to vaccinate their offspring than transmit vaccine confidence. Overall, the addition of oblique transmission to a population that would otherwise equilibrate at high vaccination coverage (**Figure S5**) leads to increased attitude transition to vaccine hesitancy and subsequently lower vaccine coverage.

**Figure 10:**
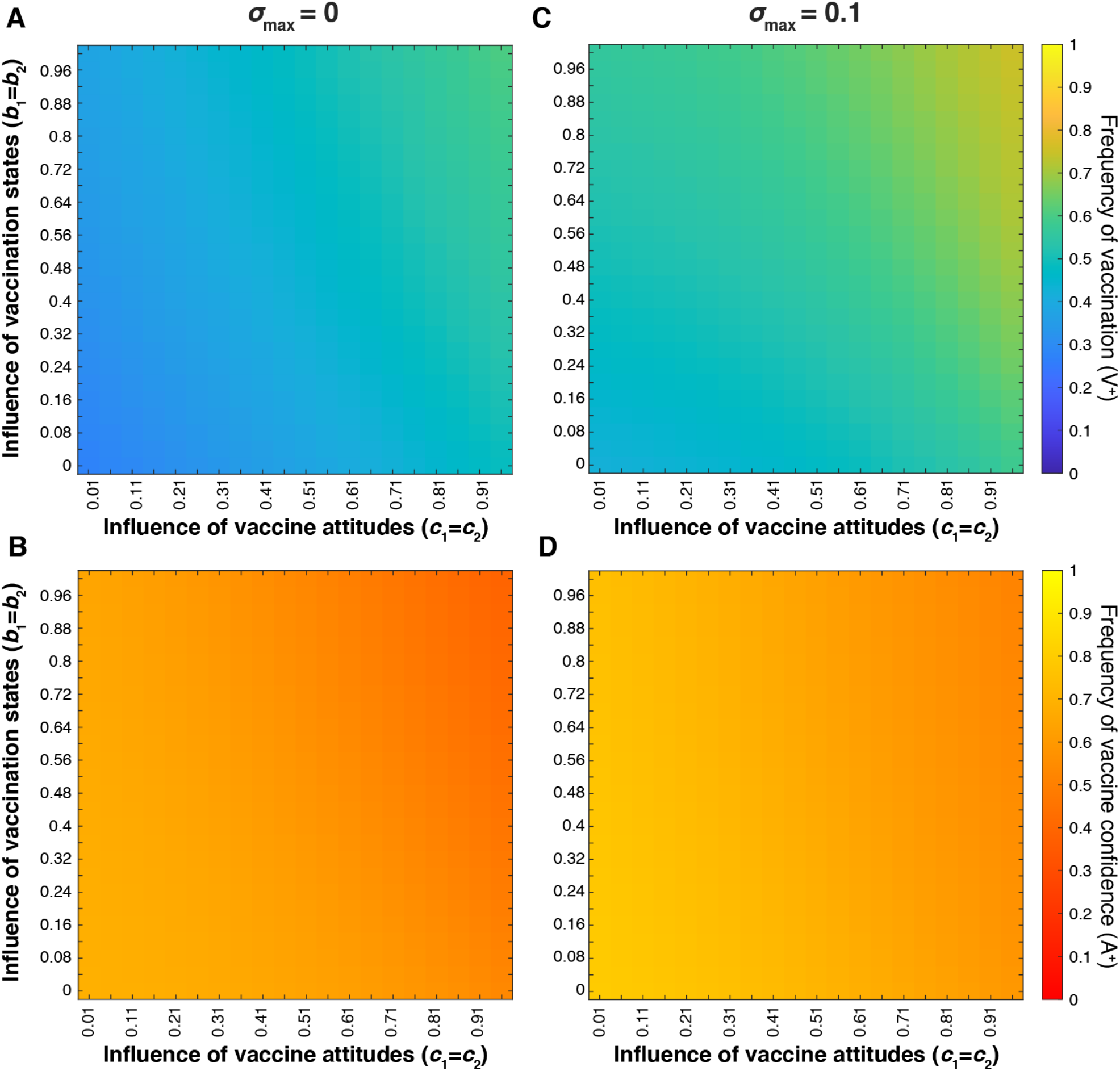
The influence of parental traits on vaccination coverage and vaccine confidence at different levels of cultural selection. Heatmaps showing final vaccination coverage (**A, C**) and final vaccination confidence **(B, D)** after 100 timesteps **with oblique transmission**. We modulate the interaction between vaccination state influence (*b*_*m*_; vertical axis) and attitude influence (*c*_*n*_; horizontal axis) at various maximum cultural selection coefficients: σ_max_ = 0 (**A, B**) and σ_max_ = 0.1 (**C, D**). As in previous figures, unvaried parameters are given in **Table 1**. Vaccination frequency increases as both influence probabilities increase and vaccination confidence decreases as both influence probabilities increase.

### 3.4 Mating Preferences

We hypothesized that mating preference (assortative mating) could modulate belief and behavior dynamics and thus the vaccination coverage and confidence levels in the population. If individuals are more likely to pair with individuals of the same vaccine attitude, such that same-attitude couples become more common and mixed-attitude couples are less common, the parameter values for mixed-attitude couples may have less impact on vaccination coverage and confidence dynamics. Therefore, we analyzed the interaction between A^+^ homophily (with *α*_1_ indicating the preference of A^+^ individuals for other A^+^ individuals) and A^−^ homophily (with *α*_2_ indicating the preference of A^−^ individuals for other A^−^ individuals) at various σ_max_ levels. When vaccine attitudes are transmitted both from parent to offspring and between unrelated individuals (vertical and oblique transmission) and there is neither cultural selection for nor against being vaccinated (σ_max_ = 0), we observe a threshold region at roughly equal mating preferences (*α*_1_ ≈ *α*_2_; diagonal lines in **Figure 11C-D**); above this boundary (when *α*_1_ > *α*_2_) vaccination coverage and confidence are much higher than below this boundary (when *α*_1_ < *α*_2_). When cultural selection explicitly does not favor vaccination (e.g. σ_max_ = −0.1, **Figure 11A-B**), low vaccination coverage and confidence can occur even when there are more vaccine confident couples in the population than hesitant couples (*α*_1_ > *α*_2_). Likewise, if cultural selection favors being vaccinated (σ_max_ > 0, **Figure 11E-H**), the threshold between high and low equilibrium values is shifted, such that high coverage and high confidence levels can potentially be attained even when vaccine hesitant individuals preferentially pair with each other more than vaccine confident individuals do (*α*_1_ < *α*_2_). We observe qualitatively similar patterns when vaccine attitudes are only transmitted from parent to offspring (**Figure S7**); as we have previously observed in **Figures 8-10**, the addition of oblique transmission leads to a broader polymorphic region than vertical transmission alone. These patterns illustrate two overarching themes: 1) preferential interactions between individuals with similar vaccine beliefs can dramatically shift the equilibrium levels of vaccination coverage and confidence with all other parameters remaining equal, and 2) the actual and perceived quality and efficacy of the vaccine are important to determining vaccine acceptance.

**Figure 11:**
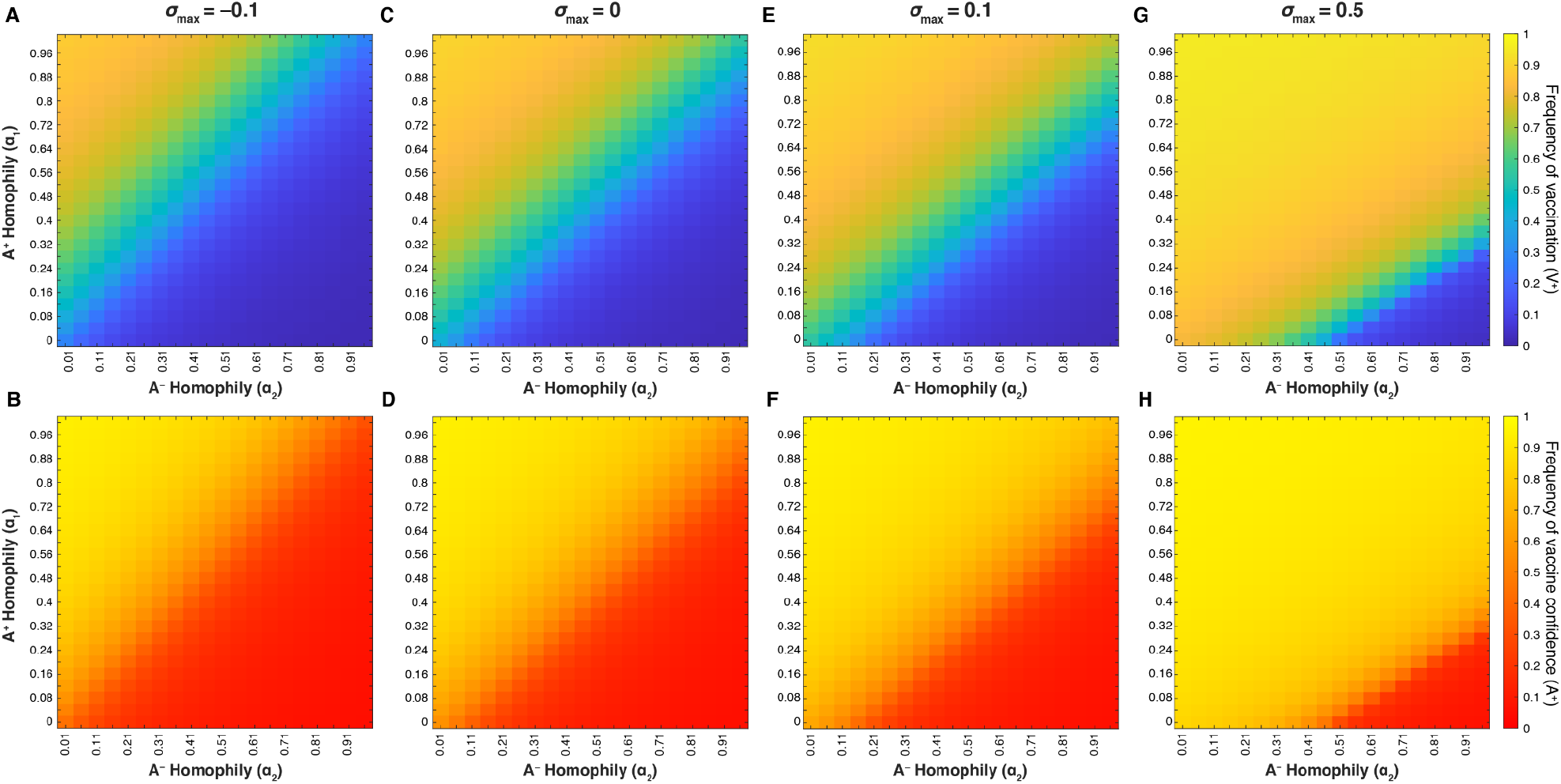
Homophily between individuals with similar vaccine beliefs can shift equilibrium frequencies of both vaccination coverage and confidence. Heatmaps showing final vaccination coverage (**A, C, E, G**) and final vaccine confidence (**B, D, F, H**) after 100 timesteps **with oblique transmission**. As in previous figures, unspecified parameters are given in **Table 1**. As vaccine-hesitant individuals (A^−^) increasingly prefer to pair with one another (*α*_2_; horizontal axis), vaccine-confident individuals (A^+^) must also preferentially interact to maintain high vaccine coverage (*α*_1_; vertical axis); this tradeoff is modulated by the cultural selection pressures on vaccination (σ_max_ = −0.1 (**A, B**), σ_max_ = 0 (**C, D**) and σ_max_ = 0.1 (**E, F**), σ_max_ = 0.5 (**G, H**)).

## 4. Discussion

In this manuscript, we present a simplified model of a complex process: the spread of vaccine attitudes and their effects on childhood vaccination frequency in a population. Increasing and maintaining sufficient vaccination coverage to combat disease is more complex than simply increasing vaccine availability or providing accurate information. A number of factors affect a person’s vaccine-related beliefs and parents’ decision to vaccinate their children, including their history with vaccinations and perception of the disease and vaccine effects. As such, it is important that we understand how these personal factors can shape vaccination cultures and thus affect public health. Using a cultural niche construction framework, we modeled the transmission of vaccine attitudes and vaccination behavior in a variety of circumstances and measured the resulting levels of vaccination coverage and vaccine confidence in the population. Using this novel approach of modeling dynamically interacting beliefs and behaviors, we are able to explore the interplay of cultural factors that drive vaccine attitudes and vaccination behavior, providing insight into how vaccination cultures are formed, maintained, and evolve.

Our model demonstrates that the cultural landscape—current policies, transmission patterns of beliefs and behaviors, etc.—can be more predictive of future levels of vaccine coverage and confidence than current coverage and confidence levels in the population. Our simulations each approached a stable equilibrium, and in general we could infer that a population with high vaccination coverage will have low rates of vaccine hesitancy and vice versa. Further, our model shows vaccine confidence transmission (*C*_*n*_) to be the parameter that most strongly determines vaccination coverage and confidence levels. That is, even though parents’ decision to vaccinate their children is based on both their level of confidence in vaccines and a consideration of their own vaccination status, the probability of transmitting vaccine-positive attitudes is a stronger predictor of future vaccination coverage than the probability of vaccination itself (**Figures 6 and 8**). Finally, our simulations also suggest that a pro-vaccination health culture can be undermined by a vaccine hesitancy “echo chamber”, possibly formed by a higher degree of preferential assortment (homophily) among vaccine-hesitant individuals, who then form pairs more likely to transmit vaccine-hesitancy to their children. Taken together, our results support the importance of considering the cultural factors that have shaped current health-related beliefs and behaviors if health policies aim to maintain or change the current conditions.

This model also shows that the perceived value and efficacy of a vaccine are important to maintaining sufficient levels of vaccination coverage, especially if vaccine confidence is not being robustly transmitted (or maintained in adulthood). Individuals essentially perform an internal cost-benefit analysis based on their circumstances and interpretation of accessible information when deciding to vaccinate. We aimed to be inclusive of their various considerations via our comprehensive cultural selection coefficient. Increasing positive public perception through honest and effective communication and reducing public concern about vaccines and increasing vaccine safety could together drive increased vaccination trust and acceptance. Achieving the optimal vaccination coverage lies not only in the hands of the public by vaccinating themselves and their children, but also in the efforts of health officials and leaders in creating an environment that fosters confidence by assuring the public of vaccine efficacy, safety, and value, while providing convenient avenues to attain vaccines.

As with any model, we cannot fully capture the complex reality of the relationship between vaccine hesitancy and vaccination behavior. First, though vaccination frequency data is available for numerous vaccines, frequency data for vaccine attitudes are much less common, with the two traditionally not surveyed together. Thus, there is no dataset that exactly estimates the phenotypes presented here, for example, the number of vaccinated but hesitant (V^+^A^−^) individuals in a population. The goals of vaccination attitude surveys have been primarily to identify themes of vaccine hesitancy, and to a lesser degree, the themes of vaccination. However, they do not report parent vaccination states or whether the child was actually vaccinated (on schedule). With data presenting parent vaccination states alongside their vaccine attitudes and vaccination decisions, we would be able to more accurately inform phenotype frequencies, possibly extending the model to incorporate various types of hesitancy. We note, however, that our results did not depend on the initial proportions of vaccination status or vaccine hesitancy, so these data would primarily be for comparison to our equilibrium outcomes.

We were also constrained by limited data to inform our cultural transmission and transition probabilities. In our model, baseline confidence transmission and influence probabilities are structured according to a simple pattern of inheritance, such that each parent is equally likely to influence an offspring’s phenotype. However, cultural traits and vaccination attitudes may not strictly follow this pattern: one parent might have more influence, or one variant of a trait might be more likely to be transmitted. In addition, transmission probabilities are constant in our model, remaining unaffected by changing cultural conditions throughout each simulation, but in reality, these probabilities may fluctuate in response to a variety of factors including vaccine type or family structure. Future developments of the model could include modulating the probability of vaccine confidence transmission according to other aspects of the cultural environment, such as the attitude frequencies in the population. We could also use the current frequency of these cultural traits across different populations to generate more specific hypotheses about their underlying cultural transmission processes [41,42] Our cultural selection coefficient and attitude transition probabilities did vary with the frequency of vaccination coverage, but the exact relationships could not be informed by existing data. Modulating both the attitude transition probabilities and the cultural selection coefficient according to the level of vaccination coverage in a population, however, reflects that perceptions about the vaccine and its associated effects on health could be meaningfully different in a population with high vaccination coverage than in one with low coverage.

Though vaccination coverage and vaccine confidence stabilized in our simulations, in reality vaccination rates fluctuate over time in response to changing population dynamics, perhaps never arriving at a stable equilibrium. For example, the increasingly rapid spread of information [43] may cause attitudes and behaviors to change frequently over short periods of time. In our model, most of the phenotype frequency fluctuations occur in the first few iterations before quickly adjusting to an equilibrium. Unlike some models of population dynamics, this model has a discrete-time format and does not consider asynchrony in population turnover. Thus, the timescale of our model might not translate directly to years or generations, and we avoid interpreting the number of iterations in literal terms. It is possible that if more realistic birth and death processes were incorporated, the cultural dynamics would occur at different timescales and would continue to fluctuate instead of approaching a stable equilibrium. In addition, the grandparents of the children to be vaccinated also influence the parents’ vaccination decisions [44]. A restructuring of the timescale or the incorporation of population asynchrony in our model could allow for consideration of these influences.

In this model, we constructed the offspring vaccination probability to be informed primarily by parents’ vaccine attitudes and secondarily by their own vaccination status. Though it is understood that there is an interaction between parents’ beliefs and their own experiences with vaccines regarding their decision to vaccinate their children, accurately modeling the relative contribution of these two factors could benefit from empirical studies on parental willingness to vaccinate based on their beliefs and vaccination status. With our current formula (*B*_*m,n*_, **Table S2**), vaccine-confident parents who did not themselves receive childhood vaccines have a reduced likelihood of vaccinating their offspring than vaccinated parents. In reality, parental vaccine attitudes might even further outweigh their own vaccination status in their decision-making process than we model here.

Finally, future developments of this model could include homophily of oblique interactions, that is, if vaccine-related beliefs influenced not only one’s mate choice but also one’s choice of social groups or information sources. On one hand, individuals who disproportionately interact with vaccine-hesitant contacts would have a biased perspective that vaccine hesitancy is more prevalent in the population than it actually is, which can reduce their likelihood of vaccinating their children [45]; on the other hand, a high degree of homophily in oblique interactions has been hypothesized to hinder the transmission of vaccine hesitancy to vaccine confident individuals, reducing the spread of the belief overall [28]. Another potential further exploration of the model includes modeling preferential assortment based on vaccination status rather than vaccine attitude, which has been shown to occur in an empirical contact-network study of high school students [46].

Our findings suggest that broad efforts to encourage and inform the public about vaccine safety and efficacy will foster higher vaccine coverage, and thus points toward several recommendations for public health policy and outreach. We recommend that accurate information about vaccines be readily accessible through a variety of means, be easily understood, and be supported by personal anecdotes since individuals who are skeptical about vaccines might invest more time in seeking out information about them [47–49], and that dialogue between people with different beliefs and attitudes be encouraged as it can help to break the “echo chambers” of homophily, encouraging individuals to communicate and empathize with one another. Therefore, to address vaccine hesitancy, our results underscore the importance of considering the cultural beliefs and influences that underpin health behaviors.

## Data Availability

All data produced will be made available online before publication.

## 5. Acknowledgements

We are grateful for feedback on this model from Glenn Webb, Ann Tate, Tony Capra, Kathy Friedman, Buddy Creech, and members of the Creanza lab. We thank the John Templeton Foundation for funding this work.

## References

1. Laland K, Matthews B, Feldman MW. An introduction to niche construction theory. Evol Ecol. 2016;30: 191–202.

2. John Odling-Smee F, Laland KN, Feldman MW. Niche Construction: The Neglected Process in Evolution (MPB-37). Princeton University Press; 2013.

3. Fogarty L, Creanza N. The niche construction of cultural complexity: interactions between innovations, population size and the environment. Philos Trans R Soc Lond B Biol Sci. 2017;372. doi:10.1098/rstb.2016.0428

4. O’Brien MJ, Laland KN, Broughton JM, Cannon MD, Fuentes A, Gerbault P, et al. Genes, culture, and agriculture: An example of human niche construction. Curr Anthropol. 2012;53: 000–000.

5. Fuentes A. Cooperation, conflict, and niche construction in the genus homo. War, peace, and human nature. 2013; 78–94.

6. Creanza N, Fogarty L, Feldman MW. Models of cultural niche construction with selection and assortative mating. PLoS One. 2012;7: e42744.

7. Creanza N, Feldman MW. Complexity in models of cultural niche construction with selection and homophily. Proc Natl Acad Sci U S A. 2014;111 Suppl 3: 10830–10837.

8. Fenner F. A successful eradication campaign. Global eradication of smallpox. Rev Infect Dis. 1982;4: 916–930.

9. Kim-Farley R, Schonberger L, Nkowane B, Kew O, Bart K, Orenstein W, et al. POLIOMYELITIS IN THE USA: VIRTUAL ELIMINATION OF DISEASE CAUSED BY WILD VIRUS. The Lancet. 1984. pp. 1315–1317. doi:10.1016/s0140-6736(84)90829-8

10. Salk D. Eradication of Poliomyelitis in the United States. I. Live Virus Vaccine-Associated and Wild Poliovirus Disease. Clinical Infectious Diseases. 1980. pp. 228–242. doi:10.1093/clinids/2.2.228

11. Thompson KM, Strebel PM, Dabbagh A, Cherian T, Cochi SL. Enabling implementation of the Global Vaccine Action Plan: developing investment cases to achieve targets for measles and rubella prevention. Vaccine. 2013;31 Suppl 2: B149–56.

12. Atwell JE, Salmon DA. Pertussis resurgence and vaccine uptake: implications for reducing vaccine hesitancy. Pediatrics. 2014. pp. 602–604.

13. Kubin L. Is There a Resurgence of Vaccine Preventable Diseases in the U.S.? Journal of Pediatric Nursing. 2019. pp. 115–118. doi:10.1016/j.pedn.2018.11.011

14. Falagas ME, Zarkadoulia E. Factors associated with suboptimal compliance to vaccinations in children in developed countries: a systematic review. Curr Med Res Opin. 2008;24: 1719–1741.

15. Dubé E, Laberge C, Guay M, Bramadat P, Roy R, Bettinger J. Vaccine hesitancy: an overview. Hum Vaccin Immunother. 2013;9: 1763–1773.

16. Scheres J, Kuszewski K. The Ten Threats to Global Health in 2018 and 2019. A welcome and informative communication of WHO to everybody. Zdrowie Publiczne i Zarządzanie. 2019. pp. 2–8. doi:10.4467/20842627oz.19.001.11297

17. Glanz JM, McClure DL, Magid DJ, Daley MF, France EK, Salmon DA, et al. Parental refusal of pertussis vaccination is associated with an increased risk of pertussis infection in children. Pediatrics. 2009;123: 1446–1451.

18. MacDonald NE, SAGE Working Group on Vaccine Hesitancy. Vaccine hesitancy: Definition, scope and determinants. Vaccine. 2015;33: 4161–4164.

19. Eggertson L. Lancet retracts 12-year-old article linking autism to MMR vaccines. CMAJ. 2010;182: E199–200.

20. Rao TSS, Andrade C. The MMR vaccine and autism: Sensation, refutation, retraction, and fraud. Indian J Psychiatry. 2011;53: 95–96.

21. Burley N. The meaning of assortative mating. Ethol Sociobiol. 1983;4: 191–203.

22. Gimelfarb A. Processes of Pair Formation Leading to Assortative Mating in Biological Populations: Encounter-Mating Model. The American Naturalist. 1988. pp. 865–884. doi:10.1086/284827

23. Salathé M, Bonhoeffer S. The effect of opinion clustering on disease outbreaks. J R Soc Interface. 2008;5: 1505–1508.

24. Perra N, Balcan D, Gonçalves B, Vespignani A. Towards a characterization of behavior-disease models. PLoS One. 2011;6: e23084.

25. Mao L, Yang Y. Coupling infectious diseases, human preventive behavior, and networks--a conceptual framework for epidemic modeling. Soc Sci Med. 2012;74: 167–175.

26. Bauch CT. Imitation dynamics predict vaccinating behaviour. Proc Biol Sci. 2005;272: 1669–1675.

27. Chauhan S, Misra OP, Dhar J. Stability analysis of SIR model with vaccination. American journal of computational and applied mathematics. 2014;4: 17–23.

28. Mehta RS, Rosenberg NA. Modelling anti-vaccine sentiment as a cultural pathogen. Evolutionary Human Sciences. 2020;2: e21.

29. May T, Silverman RD. “Clustering of exemptions” as a collective action threat to herd immunity. Vaccine. 2003;21: 1048–1051.

30. Wang E, Clymer J, Davis-Hayes C, Buttenheim A. Nonmedical exemptions from school immunization requirements: a systematic review. Am J Public Health. 2014;104: e62–84.

31. Phadke VK, Bednarczyk RA, Salmon DA, Omer SB. Association Between Vaccine Refusal and Vaccine-Preventable Diseases in the United States: A Review of Measles and Pertussis. JAMA. 2016;315: 1149–1158.

32. Pruitt RH, Kline PM, Kovaz RB. Perceived Barriers to Childhood Immunization Among Rural Populations. Journal of Community Health Nursing. 1995. pp. 65–72. doi:10.1207/s15327655jchn1202_1

33. Cavalli-Sforza LL, Feldman MW. Cultural transmission and evolution: a quantitative approach. Monogr Popul Biol. 1981;16: 1–388.

34. Bauch CT, Bhattacharyya S. Evolutionary game theory and social learning can determine how vaccine scares unfold. PLoS Comput Biol. 2012;8: e1002452.

35. Kennedy AM, Brown CJ, Gust DA. Vaccine beliefs of parents who oppose compulsory vaccination. Public Health Rep. 2005;120: 252–258.

36. Leask J. Target the fence-sitters. Nature. 2011;473: 443–445.

37. Hill HA, Singleton JA, Yankey D, Elam-Evans LD, Cassandra Pingali S, Kang Y. Vaccination Coverage by Age 24 Months Among Children Born in 2015 and 2016 — National Immunization Survey-Child, United States, 2016–2018. MMWR. Morbidity and Mortality Weekly Report. 2019. pp. 913–918. doi:10.15585/mmwr.mm6841e2

38. Hill HA, Elam-Evans LD, Yankey D, Singleton JA, Kang Y. Vaccination Coverage Among Children Aged 19–35 Months — United States, 2016. MMWR. Morbidity and Mortality Weekly Report. 2017. pp. 1171–1177. doi:10.15585/mmwr.mm6643a3

39. Gangarosa EJ, Galazka AM, Wolfe CR, Phillips LM, Gangarosa RE, Miller E, et al. Impact of anti-vaccine movements on pertussis control: the untold story. Lancet. 1998;351: 356–361.

40. Ozawa S, Mirelman A, Stack ML, Walker DG, Levine OS. Cost-effectiveness and economic benefits of vaccines in low- and middle-income countries: a systematic review. Vaccine. 2012;31: 96–108.

41. Kandler A, Powell A. Generative inference for cultural evolution. Philos Trans R Soc Lond B Biol Sci. 2018;373. doi:10.1098/rstb.2017.0056

42. Kandler A, Wilder B, Fortunato L. Inferring individual-level processes from population-level patterns in cultural evolution. R Soc Open Sci. 2017;4: 170949.

43. Hornik J, Satchi RS, Cesareo L, Pastore A. Information dissemination via electronic word-of-mouth: Good news travels fast, bad news travels faster! Computers in Human Behavior. 2015. pp. 273–280. doi:10.1016/j.chb.2014.11.008

44. Karthigesu SP, Chisholm JS, Coall DA. Do grandparents influence parents’ decision to vaccinate their children? A systematic review. Vaccine. 2018;36: 7456–7462.

45. Brunson EK. The Impact of Social Networks on Parents’ Vaccination Decisions. Immunization Strategies and Practices. 2018. pp. 123–133. doi:10.1542/9781610022774-the_impact

46. Barclay VC, Smieszek T, He J, Cao G, Rainey JJ, Gao H, et al. Positive network assortativity of influenza vaccination at a high school: implications for outbreak risk and herd immunity. PLoS One. 2014;9: e87042.

47. Gowda C, Dempsey AF. The rise (and fall?) of parental vaccine hesitancy. Hum Vaccin Immunother. 2013;9: 1755–1762.

48. Ellithorpe ME, Adams R, Aladé F. Parents’ Behaviors and Experiences Associated with Four Vaccination Behavior Groups for Childhood Vaccine Hesitancy. Matern Child Health J. 2022;26: 280–288.

49. Benin AL, Wisler-Scher DJ, Colson E, Shapiro ED, Holmboe ES. Qualitative Analysis of Mothers’ Decision-Making About Vaccines for Infants: The Importance of Trust. Pediatrics. 2006. pp. 1532–1541. doi:10.1542/peds.2005-1728

