## Supplemental figures, tables, and text for "The Cultural Evolution of Vaccine Hesitancy: Modeling the Interaction between Beliefs and Behaviors"

**Table S1. Mating frequencies for all possible matings.**

In this model, $\alpha_1$ is the rate of assortment if the choosing parent is $A^+$, and $\alpha_2$ is the rate of assortment if the choosing parent is $A^-$. The choosing parent is listed first for each mating. On the right side of the equations, the first term represents the frequency of random matings and the second term the frequency of assortative matings.

| ♂ × ♀ | Mating Frequency | ♂ × ♀ | Mating Frequency |
|---|---|---|---|
| $V^+A^+ \times V^+A^+$ | $m_{1,1} = x_1^2(1-\alpha_1) + \frac{\alpha_1 x_1^2}{(x_1+x_3)}$ | $V^-A^+ \times V^+A^+$ | $m_{3,1} = x_3x_1(1-\alpha_1) + \frac{\alpha_1 x_3 x_1}{(x_1+x_3)}$ |
| $V^+A^+ \times V^+A^-$ | $m_{1,2} = x_1x_2(1-\alpha_1)$ | $V^-A^+ \times V^+A^-$ | $m_{3,2} = x_3x_2(1-\alpha_1)$ |
| $V^+A^+ \times V^-A^+$ | $m_{1,3} = x_1x_3(1-\alpha_1) + \frac{\alpha_1 x_1 x_3}{(x_1+x_3)}$ | $V^-A^+ \times V^-A^+$ | $m_{3,3} = x_3^2(1-\alpha_1) + \frac{\alpha_1 x_3^2}{(x_1+x_3)}$ |
| $V^+A^+ \times V^-A^-$ | $m_{1,4} = x_1x_4(1-\alpha_1)$ | $V^-A^+ \times V^-A^-$ | $m_{3,4} = x_3x_4(1-\alpha_1)$ |
| $V^+A^- \times V^+A^+$ | $m_{2,1} = x_2x_1(1-\alpha_2)$ | $V^-A^- \times V^+A^+$ | $m_{4,1} = x_4x_1(1-\alpha_2)$ |
| $V^+A^- \times V^+A^-$ | $m_{2,2} = x_2^2(1-\alpha_2) + \frac{\alpha_2 x_2^2}{(x_2+x_4)}$ | $V^-A^- \times V^+A^-$ | $m_{4,2} = x_4x_2(1-\alpha_2) + \frac{\alpha_2 x_4 x_2}{(x_2+x_4)}$ |
| $V^+A^- \times V^-A^+$ | $m_{2,3} = x_2x_3(1-\alpha_2)$ | $V^-A^- \times V^-A^+$ | $m_{4,3} = x_4x_3(1-\alpha_2)$ |
| $V^+A^- \times V^-A^-$ | $m_{2,4} = x_2x_4(1-\alpha_2) + \frac{\alpha_2 x_2 x_4}{(x_2+x_4)}$ | $V^-A^- \times V^-A^-$ | $m_{4,4} = x_4^2(1-\alpha_2) + \frac{\alpha_2 x_4^2}{(x_2+x_4)}$ |

**Table S2: Probabilities of trait transmission to offspring from cultural trait pairings.**
For each mating, the probability of transmitting each trait, and corresponding influence parameters, are given. The probability of vaccinating an offspring, $B_{m,n}$, depends on both the parents' vaccination state ($V^+$: vaccinated; $V^-$: unvaccinated) and their belief state ($A^+$: vaccine confident; $A^-$: vaccine hesitant). $B_{m,n}$ is informed by the influence parameter $b_m$ that corresponds to the parents' **V** states and the influence parameter $c_n$ that correspond to their **A** states. For each parental pairing, the probability of not vaccinating an offspring is $1 – B_{m,n}$. Each pairing transmits confidence in vaccines at a rate $C_n$, and hesitancy at rate $1 – C_n$. The parameters $b_m$, $c_n$, and $C_n$ are set as constants for each simulation, and $B_{m,n}$ is calculated from these.

| | **Trait Transmission Probabilities** | | | | **Influence of parental vaccination and attitudes on offspring vaccination** | |
|---|---|---|---|---|---|---|
| **Mating pair** | **Offspring vaccination ($V^+$) probability** | **$V^-$ offspring probability** | **$A^+$ offspring probability** | **$A^-$ offspring probability** | **V influence (*m*)** | **A influence (*n*)** |
| $V^+A^+ \times V^+A^+$ | $B_{m=3,n=3} = c_3\left(\frac{1+b_3}{2}\right)$ | $1 - B_{3,3}$ | $C_3$ | $1 - C_3$ | $b_3$ | $c_3$ |
| $V^+A^+ \times V^+A^-$ | $B_{3,2} = c_2\left(\frac{1+b_3}{2}\right)$ | $1 - B_{3,2}$ | $C_2$ | $1 - C_2$ | $b_3$ | $c_2$ |
| $V^+A^- \times V^+A^+$ | $B_{3,1} = c_1\left(\frac{1+b_3}{2}\right)$ | $1 - B_{3,1}$ | $C_1$ | $1 - C_1$ | $b_3$ | $c_1$ |
| $V^+A^- \times V^+A^-$ | $B_{3,0} = c_0\left(\frac{1+b_3}{2}\right)$ | $1 - B_{3,0}$ | $C_0$ | $1 - C_0$ | $b_3$ | $c_0$ |
| $V^+A^+ \times V^-A^+$ | $B_{2,3} = c_3\left(\frac{1+b_2}{2}\right)$ | $1 - B_{2,3}$ | $C_3$ | $1 - C_3$ | $b_2$ | $c_3$ |
| $V^+A^+ \times V^-A^-$ | $B_{2,2} = c_2\left(\frac{1+b_2}{2}\right)$ | $1 - B_{2,2}$ | $C_2$ | $1 - C_2$ | $b_2$ | $c_2$ |
| $V^+A^- \times V^-A^+$ | $B_{2,1} = c_1\left(\frac{1+b_2}{2}\right)$ | $1 - B_{2,1}$ | $C_1$ | $1 - C_1$ | $b_2$ | $c_1$ |
| $V^+A^- \times V^-A^-$ | $B_{2,0} = c_0\left(\frac{1+b_2}{2}\right)$ | $1 - B_{2,0}$ | $C_0$ | $1 - C_0$ | $b_2$ | $c_0$ |
| $V^-A^+ \times V^+A^+$ | $B_{1,3} = c_3\left(\frac{1+b_1}{2}\right)$ | $1 - B_{1,3}$ | $C_3$ | $1 - C_3$ | $b_1$ | $c_3$ |
| $V^-A^+ \times V^+A^-$ | $B_{1,2} = c_2\left(\frac{1+b_1}{2}\right)$ | $1 - B_{1,2}$ | $C_2$ | $1 - C_2$ | $b_1$ | $c_2$ |
| $V^-A^- \times V^+A^+$ | $B_{1,1} = c_1\left(\frac{1+b_1}{2}\right)$ | $1 - B_{1,1}$ | $C_1$ | $1\text{-}C_1$ | $b_1$ | $c_1$ |
| $V^-A^- \times V^+A^-$ | $B_{1,0} = c_0\left(\frac{1+b_1}{2}\right)$ | $1 - B_{1,0}$ | $C_0$ | $1\text{-}C_0$ | $b_1$ | $c_0$ |
| $V^-A^+ \times V^-A^+$ | $B_{0,3} = c_3\left(\frac{1+b_0}{2}\right)$ | $1 - B_{0,3}$ | $C_3$ | $1\text{-}C_3$ | $b_0$ | $c_3$ |
| $V^-A^+ \times V^-A^-$ | $B_{0,2} = c_2\left(\frac{1+b_0}{2}\right)$ | $1 - B_{0,2}$ | $C_2$ | $1\text{-}C_2$ | $b_0$ | $c_2$ |
| $V^-A^- \times V^-A^+$ | $B_{0,1} = c_1\left(\frac{1+b_0}{2}\right)$ | $1 - B_{0,1}$ | $C_1$ | $1\text{-}C_1$ | $b_0$ | $c_1$ |
| $V^-A^- \times V^-A^-$ | $B_{0,0} = c_0\left(\frac{1+b_0}{2}\right)$ | $1 - B_{0,0}$ | $C_0$ | $1\text{-}C_0$ | $b_0$ | $c_0$ |

**Table S3: Probability range assignments for Figure 5**

To vary the range of $B_{m,n}$ used in a given simulation, each probability was grouped according to default vaccination probability calculations. All probabilities in a group hold the value assigned to that group in the range, as shown. $C_n$ probabilities were assigned values as shown, with $C_0$ taking the lowest value in the range and $C_3$ taking the highest. The lowest probability range group is given as an example of value assignment.

| | Range: Low | | → | | | High |
|---|---|---|---|---|---|---|
| Parameters | $B_{0,0}$, $B_{1,0}$, $B_{2,0}$, $B_{3,0}$ | $B_{0,1}$, $B_{0,2}$ | $B_{1,1}$, $B_{1,2}$, $B_{2,1}$, $B_{2,2}$ | $B_{3,1}$, $B_{3,2}$, $B_{0,3}$ | $B_{2,3}$, $B_{1,3}$ | $B_{3,3}$ |
| Example value (range 0–0.49) | 0 | 0.09 | 0.19 | 0.29 | 0.39 | 0.49 |

| | Low | → | | High |
|---|---|---|---|---|
| Parameters | $C_0$ | $C_1$ | $C_2$ | $C_3$ |
| Example value (range 0.1–0.4) | 0.1 | 0.2 | 0.3 | 0.4 |

**Text S1: Recursions for Vaccine Niche Construction**

$$\bar{w}x_1' = (1+\sigma_1)\big(m_{11}B_{3,3}C_3 + m_{12}B_{3,2}C_2 + m_{21}B_{3,1}C_1 + m_{13}B_{2,3}C_3 + m_{31}B_{1,3}C_3 + m_{14}B_{2,2}C_2 + m_{41}B_{1,1}C_1 + m_{22}B_{3,0}C_0 + m_{23}B_{2,1}C_1 + m_{32}B_{1,2}C_2 + m_{24}B_{2,0}C_0 + m_{42}B_{1,0}C_0 + m_{33}B_{0,3}C_3 + m_{34}B_{0,2}C_2 + m_{43}B_{0,1}C_1 + m_{44}B_{0,0}C_0\big)$$

$$\bar{w}x_2' = (1+\sigma_1)\big(m_{11}B_{3,3}(1-C_3) + m_{12}B_{3,2}(1-C_2) + m_{21}B_{3,1}(1-C_1) + m_{13}B_{2,3}(1-C_3) + m_{31}B_{1,3}(1-C_3) + m_{14}B_{2,2}(1-C_2) + m_{41}B_{1,1}(1-C_1) + m_{22}B_{3,0}(1-C_0) + m_{23}B_{2,1}(1-C_1) + m_{32}B_{1,2}(1-C_2) + m_{24}B_{2,0}(1-C_0) + m_{42}B_{1,0}(1-C_0) + m_{33}B_{0,3}(1-C_3) + m_{34}B_{0,2}(1-C_2) + m_{43}B_{0,1}(1-C_1) + m_{44}B_{0,0}(1-C_0)\big)$$

$$\bar{w}x_3' = \big(m_{11}(1-B_{3,3})C_3 + m_{12}(1-B_{3,2})C_2 + m_{21}(1-B_{3,1})C_1 + m_{13}(1-B_{2,3})C_3 + m_{31}(1-B_{1,3})C_3 + m_{14}(1-B_{2,2})C_2 + m_{41}(1-B_{1,1})C_1 + m_{22}(1-B_{3,0})C_0 + m_{23}(1-B_{2,1})C_1 + m_{32}(1-B_{1,2})C_2 + m_{24}(1-B_{2,0})C_0 + m_{42}(1-B_{1,0})C_0 + m_{33}(1-B_{0,3})C_3 + m_{34}(1-B_{0,2})C_2 + m_{43}(1-B_{0,1})C_1 + m_{44}(1-B_{0,0})C_0\big)$$

$$\bar{w}x_4' = \big(m_{11}(1-B_{3,3})(1-C_3) + m_{12}(1-B_{3,2})(1-C_2) + m_{21}(1-B_{3,1})(1-C_1) + m_{13}(1-B_{2,3})(1-C_3) + m_{31}(1-B_{1,3})(1-C_3) + m_{14}(1-B_{2,2})(1-C_2) + m_{41}(1-B_{1,1})(1-C_1) + m_{22}(1-B_{3,0})(1-C_0) + m_{23}(1-B_{2,1})(1-C_1) + m_{32}(1-B_{1,2})(1-C_2) + m_{24}(1-B_{2,0})(1-C_0) + m_{42}(1-B_{1,0})(1-C_0) + m_{33}(1-B_{0,3})(1-C_3) + m_{34}(1-B_{0,2})(1-C_2) + m_{43}(1-B_{0,1})(1-C_1) + m_{44}(1-B_{0,0})(1-C_0)\big)$$

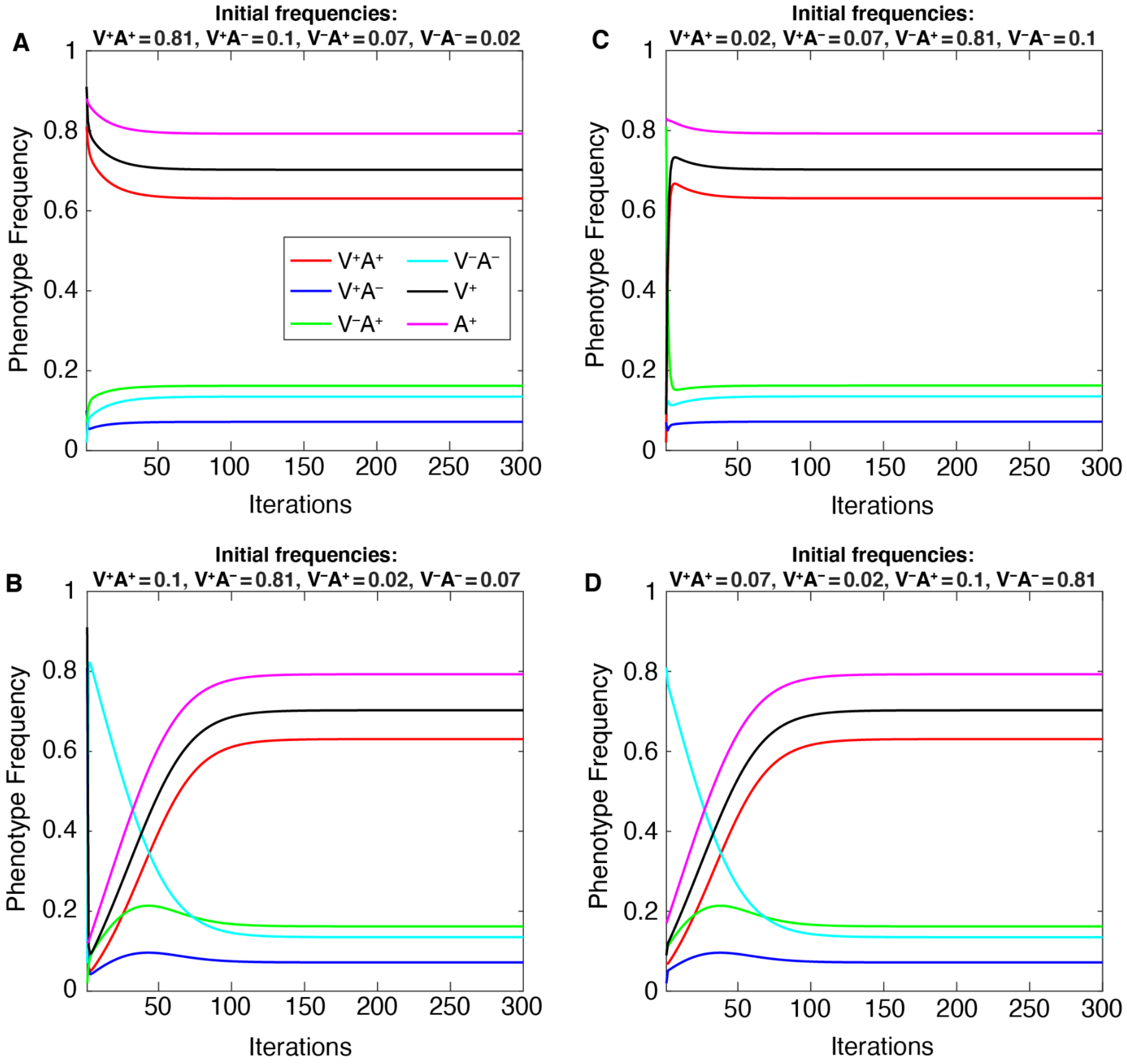


**Figure S1**: **Population frequencies reach stable equilibria determined by the parameter space, not by initial frequencies.** Each phenotype approaches the same equilibrium frequency for a given set of parameters regardless of its initial frequency in our simulations. Each of the four phenotype frequencies and the total $V^+$ and $A^+$ frequencies (vertical axis) approach equilibrium values prior to iteration 300 and remain stably at those frequencies (compare this figure with **Figure 3 (top row)**, which shows the first 100 iterations of the same simulations). We varied the initial frequencies, such that we begin each simulation with a different phenotype at an initial high frequency (0.81): **A**) $\boldsymbol{x_1}$ **($V^+A^+$) = 0.81**, $x_2$ ($V^+A^-$) = 0.1, $x_3$ ($V^-A^+$) = 0.07, $x_4$ ($V^-A^-$) = 0.02; $V^+A^-$; **B**) $x_1$ ($V^+A^+$) = 0.1, $\boldsymbol{x_2}$ **($V^+A^-$) = 0.81**, $x_3$ ($V^-A^+$) = 0.02, $x_4$ (V−A−) = 0.07; $V^-A^+$;  **C**) $x_1$ ($V^+A^+$) = 0.02, $x_2$ ($V^+A^-$) = 0.07, $\boldsymbol{x_3}$ **($V^-A^+$) = 0.81**, $x_4$ ($V^-A^-$) = 0.1 ); **D)** $x_1$ ($V^+A^+$) = 0.07, $x_2$ ($V^+A^-$) = 0.02, $x_3$ ($V^-A^+$) = 0.1, $\boldsymbol{x_4}$ **($V^-A^-$) = 0.81**. The remaining parameters are held at default values (**Table 1**) and  simulations were performed **without oblique transmission (only parent-to-offspring transmission)**. These results indicate that equilibrium frequencies are determined by the parameter space, not initial frequencies.

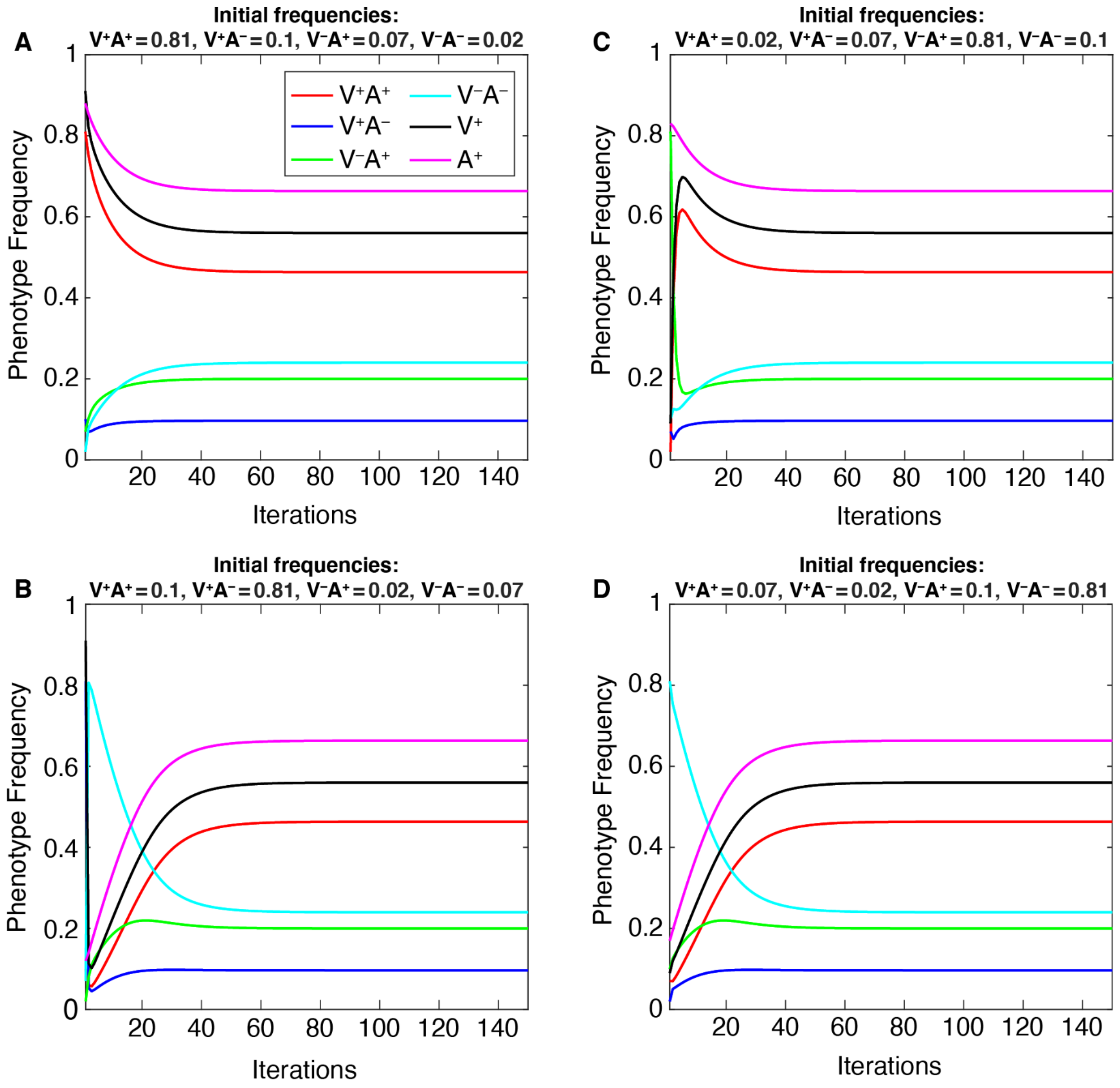


**Figure S2**: **Population frequencies reach stable equilibria determined by the parameter space, not by initial frequencies (with oblique transmission).** Each phenotype approaches the same equilibrium frequency for a given parameter set regardless of its initial frequency in our simulations. Each of the four phenotype frequencies and the total $V^+$ and $A^+$ frequencies (vertical axis) approach equilibrium values prior to iteration 300 and remain stably at those frequencies (compare this figure with **Figure 3 (top row)**, which shows results with only vertical transmission). We varied the initial frequencies, such that we begin each simulation with a different phenotype at an initial high frequency (0.81): $V^+A^+$ in panel **A**, $V^+A^-$ in panel **B**, $V^-A^+$ in panel **C**, $V^-A^-$ in panel **D**; the remaining phenotypes are set to lower frequencies (0.1, 0.07, 0.02). See **Figure S1** for a full listing of these initial frequencies. The remaining parameters are held at default values (**Table 1**) and these simulations **included both vertical and oblique transmission**.

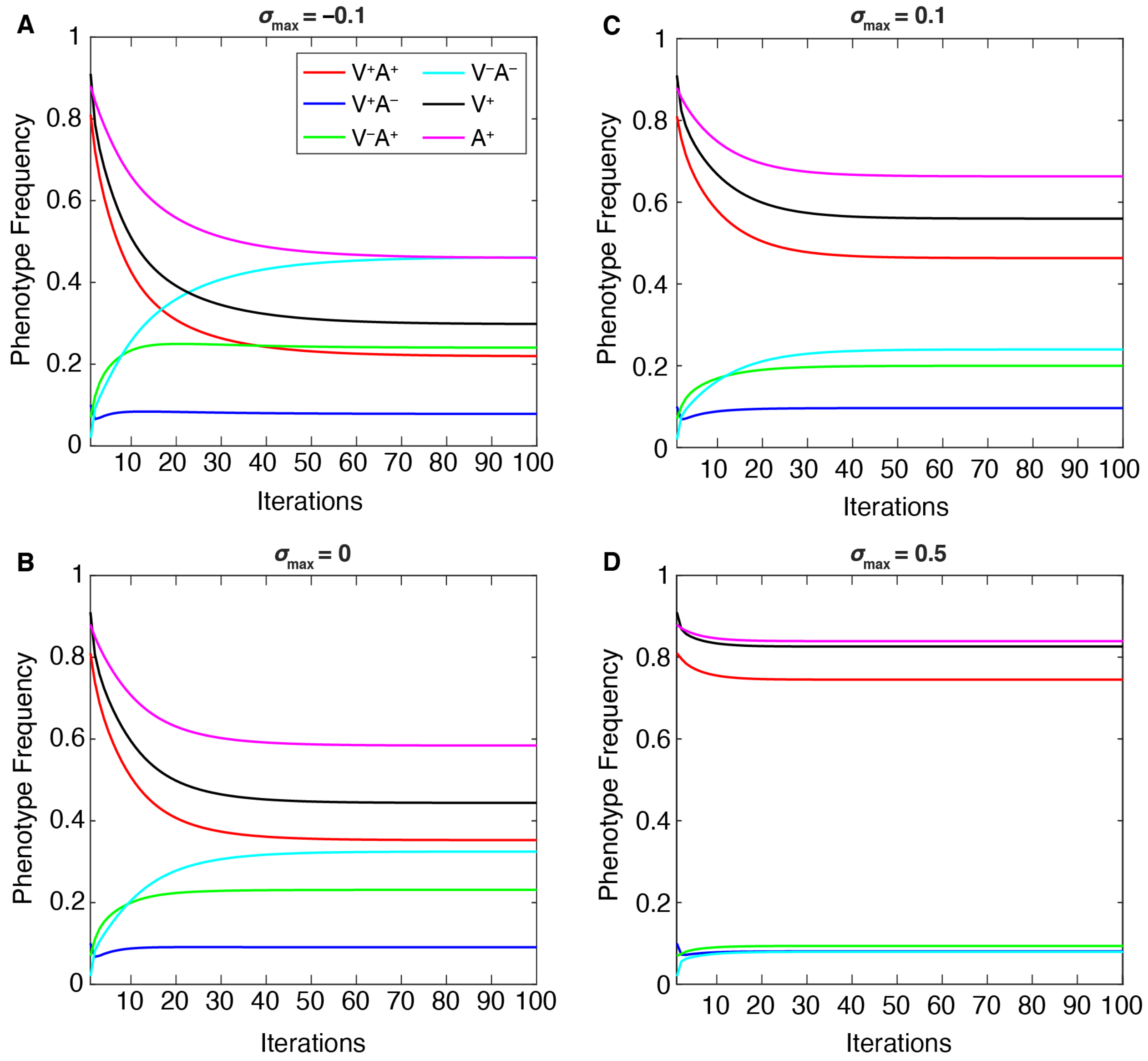


**Figure S3**: **Temporal effects of cultural selection (with oblique transmission)**. The equilibrium phenotype frequencies change as the maximum cultural selection coefficient ($\sigma_{max}$) is varied: **A.** $\sigma_{max}$ = –0.1; **B.** $\sigma_{max}$ = 0; **C.** $\sigma_{max}$ = 0.1; **D.** $\sigma_{max}$ = 0.5. Oblique transmission is included, and other parameters are held at default values (**Table 1**). (Compare this figure with **Figure 3 (bottom row)**, which shows results with vertical transmission only.) Cultural selection against vaccinated individuals increases the frequency of $V^-A^-$, while decreasing the other frequencies (**A**), whereas increased cultural selection favoring vaccinated individuals increases $V^+A^+$ frequencies while decreasing the other frequencies (**C**, **D**). The highest levels of conflicting phenotypes ($V^+A^-$ and $V^-A^+$) were observed when cultural selection was neutral (**B**).

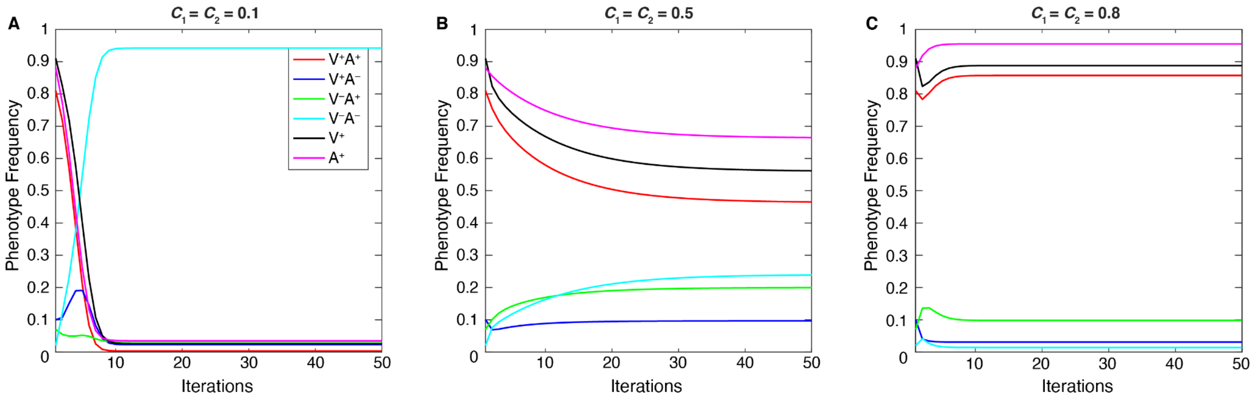


**Figure S4**: **Temporal effects of confidence transmission (with oblique transmission)**. The change in phenotype frequencies over 50 iterations as vaccine confidence transmission in mixed couples ($C_1$=$C_2$) is varied (**A**. $C_1$=$C_2$=0.1; **B**. $C_1$=$C_2$=0.5; **C**. $C_1$=$C_2$ = 0.8). Oblique transmission is included and other parameters are held at default values (Table 1). The population equilibrates at over 90% $V^-A^-$ at low confidence transmission (**A**). Increasing the probability of confidence transmission results in higher $V^+A^+$ frequencies and lower $V^-A^-$ (**B**, **C**). In comparison to simulations with only vertical transmission (**Figure 4**), phenotype frequencies reach equilibrium at values closer to mid-range levels (i.e. $V^+A^+$ levels at equilibrium are reduced while other frequencies are increased). It is also worth noting that the $V^-A^-$ equilibrium value is higher than $V^-A^+$ at $C_1$=$C_2$ = 0.5 when oblique transmission is added.

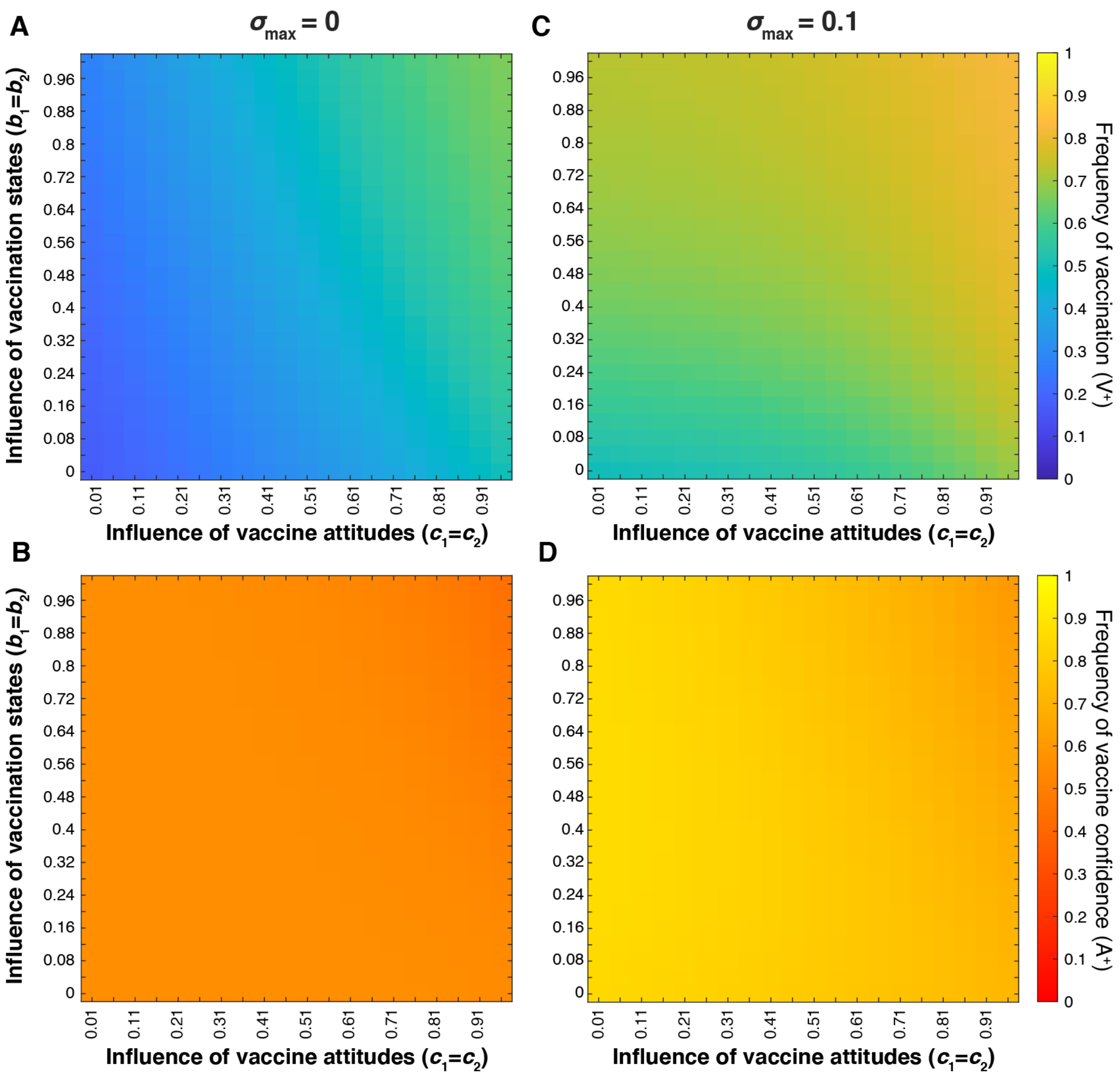


**Figure S5**: **The influence of parental traits on vaccination coverage and vaccine confidence (without oblique transmission).** Equilibrium vaccination coverage (**A,C**) and corresponding vaccine confidence (**B, C**) after 100 time-steps without oblique transmission– only parent-to-offspring transmission (Compare to **Figure 10** with oblique transmission). Influence of parental vaccination states ($b_1$ =$b_2$; vertical axis) and influence of parent vaccine attitudes ($c_1$ =$c_2$; horizontal axis) are varied at two maximum cultural selection coefficients: $\sigma_{max}$= 0 (**A, B**) and $\sigma_{max}$= 0.1 (**C, D**). Positive selection for vaccination increased vaccination coverage and vaccine confidence across parameter ranges, however, vaccine confidence is lower than expected at the intersection of high state influence parameters.

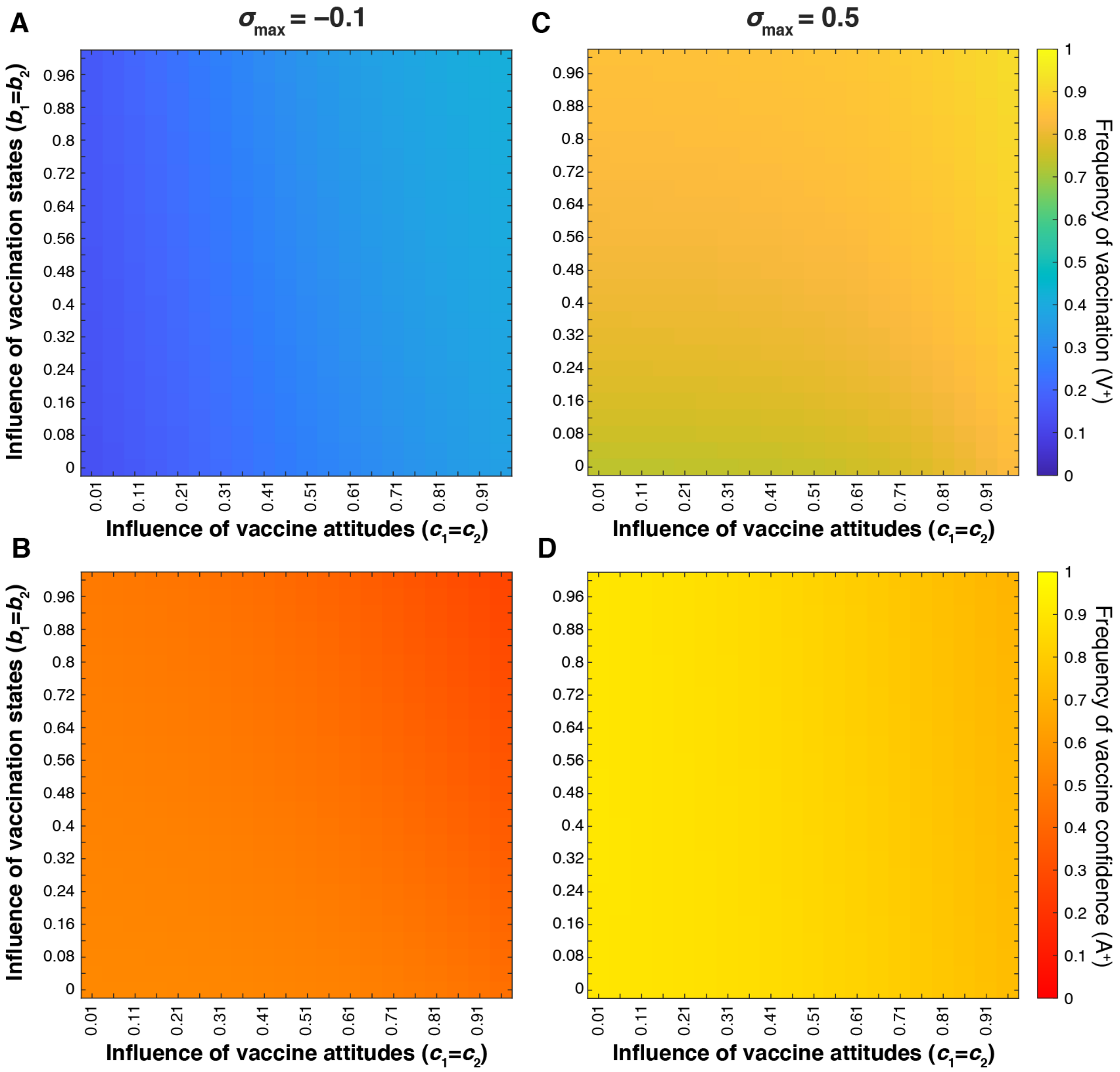


**Figure S6**: **The influence of parental traits on vaccination coverage and vaccine confidence (at extreme levels of cultural selection compared to Figure 10)**. Equilibrium vaccination coverage (**A,C**) and corresponding vaccine confidence (**B, C** respectively) after 100 time-steps **with oblique transmission**. Influence of parental vaccination states ($b_1 = b_2$; vertical axis) and influence of parent vaccine attitudes ($c_1 = c_2$; horizontal axis) are varied at two maximum cultural selection coefficients: $\sigma_{max}$= -0.1 (**A, B**) and $\sigma_{max}$= 0.5 (**C, D**). Positive selection for vaccination increases vaccination coverage and vaccine confidence across parameter ranges, however, vaccine confidence is lower than expected at the intersection of high state influence parameters.

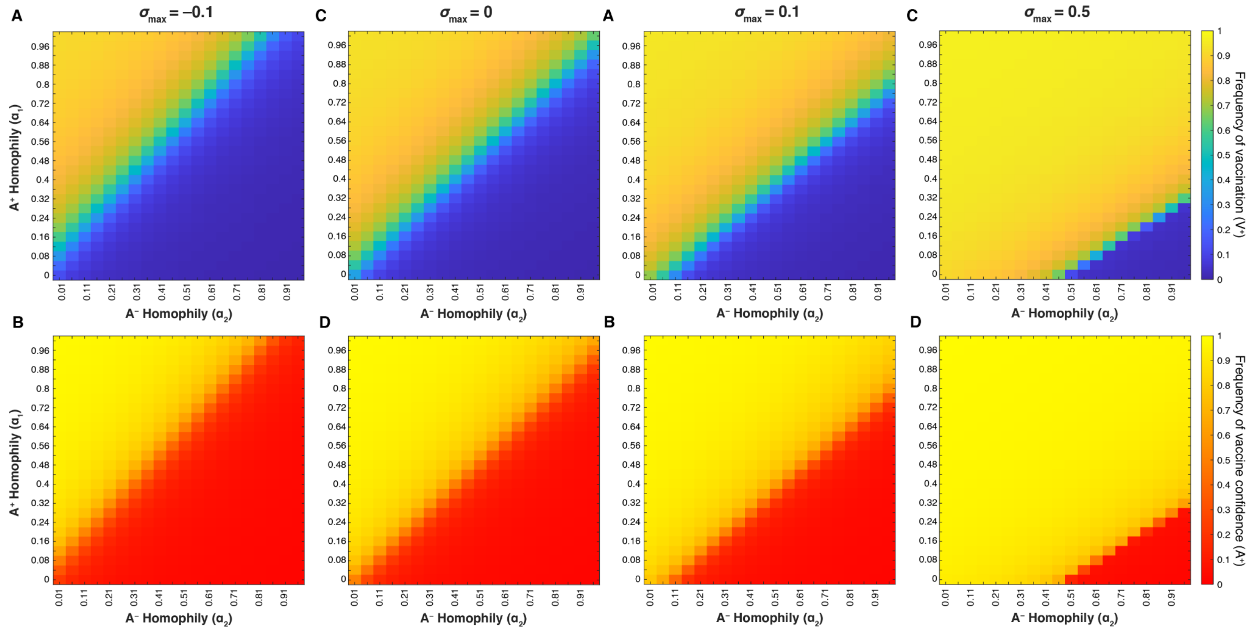


**Figure S7: Homophily between individuals with similar vaccine beliefs can shift the equilibrium frequencies of both vaccination coverage and confidence (without oblique transmission).** Heatmaps showing final vaccination coverage **(A, C)** and final vaccine confidence **(B, D)** after 100 timesteps without oblique transmission (only parent-to-offspring transmission; compare to **Figure 11**). As in previous figures, unspecified parameters are given in **Table 1**. As vaccine-hesitant individuals ($A^-$) increasingly prefer to pair with one another (increasing $\alpha_2$; horizontal axis), vaccine-confident individuals ($A^+$) must also preferentially interact to maintain high vaccine coverage ($\alpha_1$; vertical axis); this tradeoff is modulated by the cultural selection pressures on vaccination ($\sigma_{max}$ = -0.1 (**A, B**), $\sigma_{max}$ = 0 (**C, D**), $\sigma_{max}$ = 0.1 (**E, F**) and $\sigma_{max}$ = 0.5 (**G, H**)).
